# Sociodemographic Characteristics and Longitudinal Progression of Multimorbidity: A Multistate Modelling Analysis of a Large Primary Care Records Dataset in England

**DOI:** 10.1101/2023.03.06.23286491

**Authors:** Sida Chen, Tom Marshall, Christopher Jackson, Jennifer Cooper, Francesca Crowe, Krish Nirantharakumar, Catherine L Saunders, Paul Kirk, Sylvia Richardson, Duncan Edwards, Simon Griffin, Christopher Yau, Jessica K Barrett

**Affiliations:** MRC Biostatistics Unit, University of Cambridge; Institute of Applied Health Research, University of Birmingham; Department of Public Health and Primary Care, University of Cambridge; Nuffield Department for Women’s & Reproductive Health, University of Oxford and Health Data Research UK

## Abstract

**Background:** Multimorbidity, characterized by the coexistence of multiple chronic conditions in an individual, is a rising public health concern. While much of the existing research has focused on cross-sectional patterns of multimorbidity, there remains a need to better understand the longitudinal accumulation of diseases. This includes examining the associations between important sociodemographic characteristics and the rate of progression of chronic conditions.

**Methods and findings:** We utilized electronic primary care records from 13 million participants in England, drawn from the Clinical Practice Research Datalink (CPRD Aurum), spanning from 2005 to 2020 with a median follow-up of 4.71 years (IQR: 1.78, 11.28). The study focused on five important chronic conditions: cardiovascular disease, type-2 diabetes, chronic kidney disease, heart failure and mental health conditions. Key sociodemographic characteristics considered include ethnicity, social and material deprivation, gender, and age. We employed a flexible spline-based parametric multistate model to investigate the associations between these sociodemographic characteristics and the rate of different disease transitions throughout multimorbidity development. Our findings reveal distinct association patterns across different disease transition types.

Deprivation, gender and age generally demonstrated stronger associations with disease diagnosis compared to ethnic group differences. Notably, the impact of these factors tended to attenuate with an increase in the number of pre-existing conditions, especially for deprivation, gender and age. Furthermore, the impact of deprivation, gender, and age was typically more pronounced when transitioning from a mental health condition. A primary limitation of our study is that potential diagnostic inaccuracies in primary care records, such as underdiagnosis, overdiagnosis, or ascertainment bias of chronic conditions, could influence our results.

**Conclusion:** Our results indicate that early phases of multimorbidity development could warrant increased attention. The potential importance of earlier detection and intervention of chronic conditions is underscored, particularly for mental health conditions and higher-risk populations. These insights may have important implications for the management of multimorbidity.

**Author summary:** *Why Was This Study Done?:*

- Multimorbidity, the presence of two or more chronic conditions in an individual, is a growing concern in aging societies. A better understanding of how these conditions develop and progress over time, and the factors associated with this process, is important for more effective management and treatment.
- Previous research has analysed the association between certain socioeconomic and behavioural factors and the rate of disease progression over time. However, these studies typically focused on a limited number of conditions and rarely considered all possible combinations. Furthermore, their analyses often rely on relatively small datasets.
- There is a gap in our detailed understanding of the impact of sociodemographic characteristics – such as ethnicity, deprivation, age, and gender - on the progression of multiple chronic conditions.

*What Did the Researchers Do and Find?:*

- We analysed the health records of over 13 million participants in England from 2005 to 2020, focusing on how factors like ethnicity, deprivation, gender and age are associated with the accumulation of five common conditions: cardiovascular disease (CVD), type-2 diabetes (T2D), chronic kidney disease (CKD), heart failure (HF), and mental health conditions (MH).
- We found that factors like deprivation, age, and gender generally have a stronger link to the diagnosis of these conditions compared to ethnicity. Moreover, the impact of deprivation, age and gender tend to be weakened as the number of pre-existing conditions a person has increases.
- In particular, when an individual already has a mental health condition, and if they were older, male, or from more deprived groups, they were expected to develop other conditions like CVD, T2D and HF more quickly compared to scenarios involving other pre-existing conditions.

*What Do These Findings Mean?:*

- ur findings suggest that early stages, when people are starting to develop multiple health issues, especially when mental health problems are first diagnosed and in high-risk groups, may require more attention for improved patient care and healthcare strategies.
- ur results underscore the need to investigate and better understand the different biological, psychological, and societal factors that influence the progression to multimorbidity.
- that our analysis is based on health records, which may have incomplete or inaccurate information, including potential inaccuracies in condition diagnosis. These limitations may have an influence on our results.

## Introduction

Multimorbidity or multiple long-term conditions is defined as the co-existence of two or more chronic conditions in an individual [1]. Multimorbidity increases with age and because of the complexity of managing those with multiple health conditions, it is of increasing importance in countries with ageing populations [2]. Existing research indicates that multimorbidity is associated with poorer quality of life, worsened health outcomes, increased healthcare use and higher mortality [3,4]. Multimorbidity develops sequentially, as individuals accumulate diagnoses, but to date, research has mainly described cross-sectional patterns of multimorbidity, primarily focusing on estimating multimorbidity prevalence and identifying multimorbidity clusters [5–8]. We need to understand how diseases accumulate over time and the associations of potentially important sociodemographic characteristics with this accumulation process. This may help identify those at high risk of developing further diseases. A recent systematic scoping review summarised research on longitudinal modelling and analysis of multimorbidity patterns, see [9]. One of these analyses investigated the sequence of occurrence of diseases using a visualisation approach, stratified by deprivation status, age and ethnicity [10]. They reported that the more socially deprived had a greater risk for multimorbidity and that ethnicity was associated with the development of multimorbidity.

The progression of multimorbidity can be conceptualized as an individual transitioning through various states of comorbidity, each representing a distinct disease profile. This sequence of disease accumulation can be viewed as a specific form of event history. Multistate models (MSM) extend the traditional survival model to encompass more than two states, providing a useful probabilistic framework for modelling complex longitudinal event history data and making predictive inferences. MSMs have been effectively used in recent multimorbidity studies. For instance, MSMs have been applied to investigate the progression of three chronic conditions in an elderly population [11], and to analyse socioeconomic inequalities in life expectancy with and without multimorbidity [12]. They have also been used to assess the influence of lifestyle factors on disease accumulation [13], and to explore the effects of sociodemographic and clinical factors on several stages of multimorbidity development [14]. Other studies have utilised MSMs to examine the potentially differing impacts of lifestyle risk factors on cardiometabolic multimorbidity [15], and to assess the role of midlife factors on different cardiometabolic disease trajectories [16]. In a more recent study, a 17-state MSM was used to examine the impact of the order of disease acquisition on patient mortality [17].

Despite the significant potential of MSMs for multimorbidity analysis, their current utilization is often constrained by computational challenges associated with model estimation and/or constraints in sample size. Most existing MSMs for multimorbidity have been restricted to incorporate only a minimal number of conditions (e.g. three), or represent multimorbidity status merely as counts of conditions rather than discerning specific condition combinations to simplify the model structure. In addition, current models largely rely on simple parametric models such as the Weibull hazard model, which has limited flexibility, or Cox semi-parametric models, which are incapable of absolute risk quantifications or predictions over time. While certain risk factors such as age, gender and some lifestyle factors have been considered, detailed studies on the impact of sociodemographic characteristics on the rate of disease-specific multimorbidity progression remain scarce. Furthermore, the reliance on relatively small cohort samples in most of the previous analyses may limit the power of their findings relative to the number of states of the models and subgroups.

In this work, we propose a flexible parametric multistate modelling framework for describing the temporal progression of five chronic conditions and investigating the association between several key sociodemographic risk factors, including age, gender, ethnicity and social and material deprivation, and the rate of different disease transitions. We focus on cardiovascular disease (CVD), type 2 diabetes (T2D), chronic kidney disease (CKD), mental health conditions (MH) and heart failure (HF), which are cardiometabolic, renal conditions and mental conditions that are common, important, and well-coded in primary care due to the Quality and Outcomes Framework (QoF) incentivisation program for general practice. These conditions have well-established treatment and prevention strategies that are known to frequently co-occur, especially in socio-economically deprived areas [18]. By treating different disease combinations as separate states, our model comprises a total of 33 states and 112 transitions and is estimated based on a large dataset of approximately 13 million English electronic primary care records.

Importantly, our MSM extends previous works by allowing for more comorbidity states and disease transition types, thus providing a more detailed characterization of the impacts of the sociodemographic factors on the progression of these five diseases. While the framework is flexible and capable of accommodating different or additional conditions in principle, our primary analysis centers on these five conditions to avoid overcomplicating the presentation and interpretation of results.

## Methods

### Ethics statement

The Clinical Practice Research Datalink (CPRD) Research Database has ethics approval from the Health Research Authority to support research using anonymised patient data (East Midlands - Derby Research Ethics Committee, reference 21/EM/0265). Data was approved for this study by the CPRD Independent Scientific Advisory Committee (protocol 19_265).

### Data sources

The study investigated the temporal sequence of diagnosed chronic conditions in a large anonymised dataset of electronic primary care records, the Clinical Practice Research Datalink (CPRD) [19]. The CPRD Aurum database contains routinely collected primary care electronic health records from 19 million participants across the 738 English general practices that use EMIS Web® software [19] (10% of practices and 13% of the English population). It is representative of the English population in terms of age, gender, regional geographical spread and deprivation. It contains information on individual participant demographics. Clinical observations, diagnoses, test results, immunisations, lifestyle factors, and prescriptions are recorded as Read Version 2, SNOMED-CT, and EMIS Web® clinical codes. Data are available from 1995 onwards, and there is a median follow-up of 4.2 years (IQR: 1.5 - 11.4) for all patients and 9.1 years (IQR: 3.3 - 20.1) for currently registered patients [19].

### Conditions

In this study, we placed our attention on five specific chronic conditions: 1) cardiovascular disease (CVD), which comprises ischaemic heart disease (IHD) and stroke or transient ischaemic attack (TIA); 2) type-2 diabetes (T2D); 3) chronic kidney disease (CKD); 4) mental health conditions (MH), comprising depression and/or anxiety; and 5) heart failure (HF). Within CVD, we distinguished between IHD, which primarily affects the heart, and stroke or TIA, which primarily affects the brain. For mental health conditions, we concentrated on depression and anxiety as they are the most prevalent mental health conditions [20], and, while there is clinical overlap in their presentation, they are typically phenotypically quite distinct from other less common mental health conditions such as psychotic disorders, post-traumatic stress disorder and substance misuse disorders. The presence of a chronic condition was determined by the occurrence of one of the relevant clinical codes (code lists are provided in the supplementary material) in the participant records (based on primary care data only), and the date of diagnosis was taken as the date of the first occurrence of any associated clinical codes. An absence of the code was considered to indicate an absence of the respective condition.

### Sociodemographic characteristics

Sociodemographic characteristics include ethnicity, social and material deprivation, age (calculated from the year of birth) and sex (male/female). Ethnicity was initially classified into five different groups, including White, South Asian (South Asian or British Asian), Black (Black African, Black Caribbean or Black British), Mixed and Others (ethnicity codings are provided in the supplementary material) [21]. In our multistate analysis described below, Mixed was merged into Others due to their infrequency in many disease transitions. Social and material deprivation were approximated using the English Index of Multiple Deprivation (IMD), which is derived from a range of factors including income, employment, education, crime, and living environment. The IMD is a widely accepted, area-based composite measure of area-level deprivation [22]. In the raw CPRD Aurum data, each participant was assigned an IMD decile, ranging from 1

(least deprived) to 10 (most deprived), based on their area of residence. For our analysis, we consolidated IMD deciles into quintiles to facilitate the presentation of our results. This level of granularity seemed sufficient for our purpose, as we are primarily interested in the general association patterns between deprivation levels and disease progression.

### Cohort description

Participants were eligible for inclusion in the study if they were registered in a general practice contributing to CPRD between the study start date and the study end date. The study start date was defined as the latest of: the date the participant turned 18, 01/01/2005, or six months after the participant registration start date. The study end date was defined as the earliest of the participant registration end date, date of death, or the last collection date for practice (11/05/2020). The minimum period of registration ensures time for participant characteristics to be recorded and prior diagnoses to be transferred/recorded.

Participants were followed up continuously during the study period. For conditions where code lists were combined the earliest valid code in any code list was used as the date of incidence, and long-term conditions recorded before the study start date were treated as prevalent. In preparing our dataset for analysis, we began by excluding participants aged over 90 years at the study start date (n=194,726). From the remaining participants, we further excluded those with gender classified as indeterminate (n=395), those with missing data on IMD (n=40,564), and those with missing ethnicity information (n=6,345,105). Additionally, participants with no follow-up period (n=1,199,267) were removed from the dataset. The final cohort consists of a total of 13.48 million participants. More details regarding the sample selection procedure are provided in supplemental Figure A.

### Statistical method

To evaluate how sociodemographic characteristics such as ethnicity, deprivation, age, and sex are associated with the rate of different disease accumulation during multimorbidity development, we considered a MSM with a total of 33 states and 112 allowable direct transitions (see Figure 1 for an illustration). The states incorporate all possible combinations of the five conditions a person may have during the process of disease progression and the permitted instantaneous state-to-state transitions were indicated by a direct solid line with an arrow indicating the direction of the transition. The death state is the terminal state and can be directly reached from any other state. There are two simplifying assumptions underlying the multistate diagram shown in Figure 1. The first assumption is that once a chronic condition is diagnosed, it remains ‘switched on’. This practice is common in large-scale health research and is also necessitated by the limitations of our available data. The second assumption is that no more than one event per person (either a diagnosis or death) can happen on the same day. This is based on the observation that ‘simultaneous transitions’ – two or more events happening on the same day – are extremely rare. Among our cohort of 13.47 million participants, the most frequent instances of such transitions didn’t exceed about 5000 cases (i.e. less than 0.04%). In fact, these same-day diagnoses are likely to be artificially increased by same-day diagnostic tests or recording of diagnoses, which raises questions about the accuracy of the timing of these recorded transitions. Therefore, we excluded the data associated with these simultaneous transitions from our analysis.

**Figure 1.**
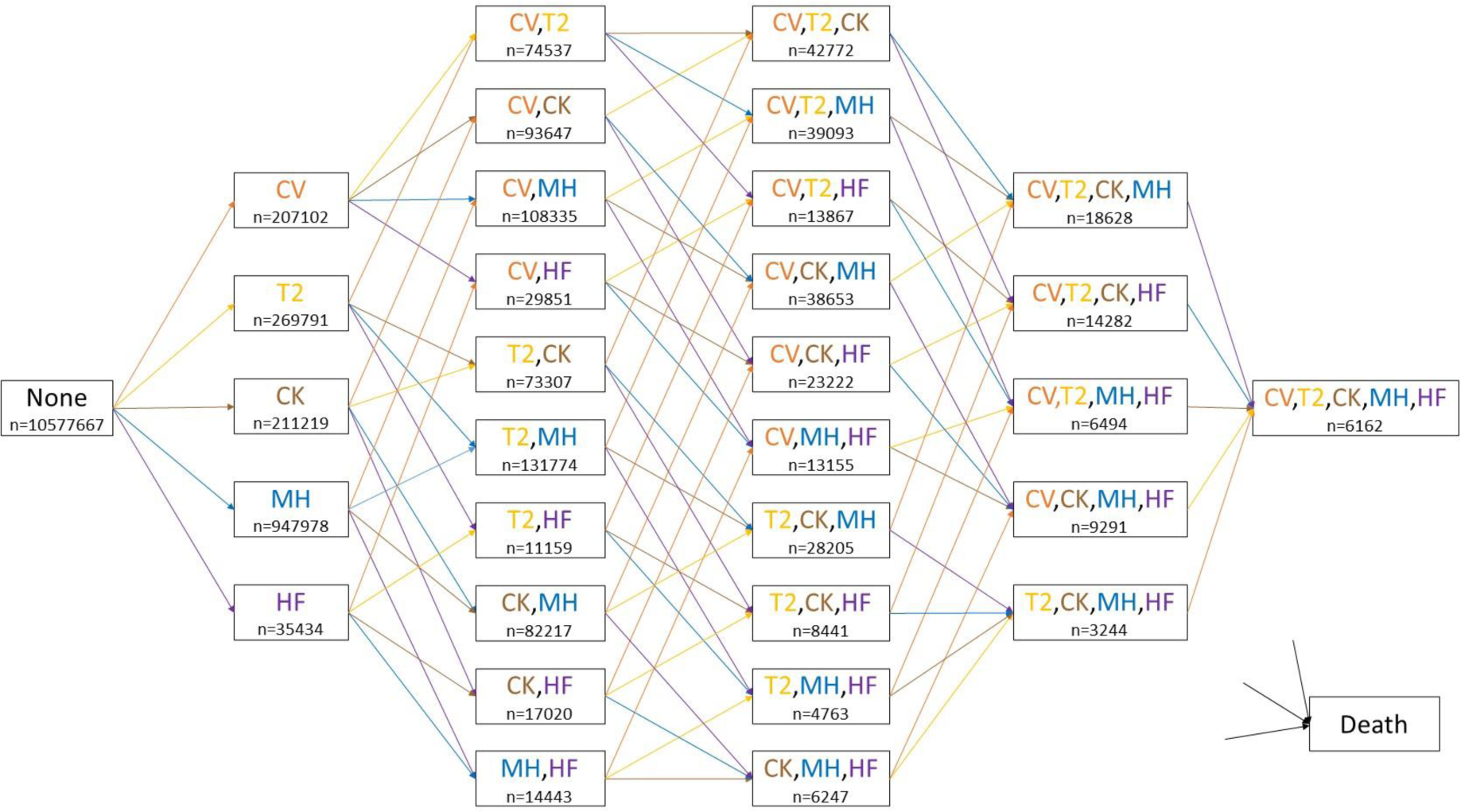
The multistate diagram depicting disease progression. The initial state, labelled “None”, represents participants with no comorbidities at the start of the study. Intermediate comorbidity states are color-coded as follows: cardiovascular disease (CV, orange), type-2 diabetes (T2, yellow), chronic kidney disease (CK, brown), mental health conditions (MH, blue), and heart failure (HF, purple). Each colored arrow indicates a transition, with the colour corresponding to the disease that is diagnosed when the transition takes place (e.g. orange arrows indicate transitions into a CVD diagnosis). The terminal state, “Death”, is directly reachable from all other states. The ‘n’ below each comorbidity state denotes the total number of participants who entered that state during the study period.

To flexibly specify the MSM, we followed the suggestions of [23] to use a separate time-to-event model for each transition, and we used a clock-reset time scale (i.e. the time scale resets to zero after every transition) so that the outcome of interest is the time from one disease diagnosis to the next diagnosis or death. More specifically, we modelled the logarithm of the cumulative transition-specific intensities as a spline function of log time [24]. This allowed us to estimate hazard ratios (HR) for each factor - age, sex, ethnicity and deprivation - adjusted for the other factors (e.g. HRs for the ethnic group are adjusted for age, sex and IMD). To limit the model’s complexity and facilitate summarization and consistent interpretation of results, we assumed proportional hazards and no interactions between these predictors for each transition. When fitting each transition, we considered data from all participants who underwent that transition during the study period. Any diagnoses that occurred before the study’s start date were treated as prevalent. Model parameters were estimated and their uncertainties quantified based on maximum likelihood estimation (MLE) theory. We were able to do this efficiently by leveraging the factorization property of the model likelihood, which allows us to fit each transition independently, and thus parallel computing could be exploited; see ‘Supplemental technical details’ section for additional details. In our implementation, we utilised the flexsurv package [25] within the R software environment. These computations were performed on the Cambridge Service for Data Driven Discovery (CSD3) High-Performance Computing (HPC) system using the Ice Lake CPUs.

To investigate the potential association between the number of pre-existing conditions and the hazard ratios associated with each sociodemographic characteristic for transitions into each of the five conditions, we further conducted a meta-regression analysis based on the fitted MSM. The dependent variable in our meta-regression is the estimated log-transformed hazard ratio(s) for a given characteristic, pertaining to a specific transition, and the independent variable is the number of pre-existing conditions prior to the corresponding transition. To account for heterogeneity among various types of transitions, we incorporated a random effect into the model. The analysis was implemented using the metafor R package [26].

To assess the robustness of our results under various modified settings, we conducted a series of sensitivity analyses. First, we evaluated the impact of condition settings on the results. We substituted HF, which is more similar to CVD among the five conditions, with another condition that affects a different organ system - here we choose chronic obstructive pulmonary disease (COPD). Subsequently, we expanded the multistate transition network by incorporating COPD as the sixth condition, in addition to the five selected conditions. We also evaluated the impact of condition definitions, which are relevant to CVD and MH conditions as they are a composite of multiple sub-conditions. We considered the following four settings with modified definitions of CVD or MH. For CVD, we considered narrower definitions based on: 1) conditions specifically affecting the brain, i.e. comprising only stroke or TIA codes; 2) conditions specifically affecting the heart, i.e. including only the IHD code. Similarly, for MH, we considered narrower definitions based on: 1) anxiety only or 2) depression only. To assess the impact of changing the resolution of deprivation groups, we considered fitting each transition using the original IMD deciles. Additionally, we examined the impact of data exclusion on the inferences (here we focused on missing ethnicity as it accounts for the vast majority of missing data). We expanded our cohort to include participants with missing ethnicity information, assigning them to an artificial ethnic group labelled “missing”. In each scenario considered above, we re-fitted the model following the same procedure described above and examined the estimated relationships between sociodemographic variables and disease transitions. Lastly, to ensure our results are robust across different modelling approaches, we considered an alternative popular approach by specifying the MSM using transition-specific semi-parametric Cox models. HRs for each transition were estimated in a parallel fashion as above using the ‘survival’ package [27] in R.

## Results

### Summary statistics

The cohort baseline characteristics are shown in Table 1. At study entry, 78.5% (n=10,577,667) of the eligible participants had no diagnosis of any of the five conditions of interest. By the end of the follow-up, 83.7% (n=8,856,098) of these had not developed any of the five conditions, 13.8% (n=1,462,966) had developed one, 2% (n=209,237) had developed two, 0.4% (n=42504) had developed three, 0.06% (n=6399) four and 0.0044% (n=463) five of the conditions. Five percent of the study population (n=670,753) died during the study period. Detailed summary statistics for each comorbidity state in our MSM are presented in Supplementary Table A. A summary of the incidence rate for each disease transition in our MSM is provided in Supplementary Table B. The incidence rates of transitions to the first morbidity were notably lower compared to the other transitions, and generally, the incidence rates for transitions increased with the number of comorbidities. It should be noted that the relatively lower incidence rates for transitions from MH compared to other disease transitions could be due to the fact that patients in the MH state were generally younger (this age adjustment is incorporated into our MSM analysis).

**Table 1.** Baseline sociodemographic characteristics of study participants.

|  |  |  |
| --- | --- | --- |
| Cohort population |  | 13,476,121 |
| Sex (n (%)) | Male | 6,315,448 (46.9%) |
|  | Female | 7,160,673 (53.1%) |
| Age at cohort entry (median [IQR]) |  | 35 [25, 51] |
| Ethnicity (n (%)) | White | 10,753,663 (79.8%) |
|  | South Asian | 1,460,750 (10.8%) |
|  | Black | 741,504 (5.5%) |
|  | Mixed | 257,651 (1.9%) |
|  | Others | 262,553 (1.9%) |
| Index of multiple deprivation (n (%)) | 1 - least deprived | 2,384,659 (17.7%) |
|  | 2 | 2,530,868 (18.8%) |
|  | 3 | 2,620,501 (19.4%) |
|  | 4 | 3,139,349 (23.3%) |
|  | 5 - most deprived | 2,800,744 (20.8%) |
| Follow-up period (median (IQR)) |  | 4.71 [1.78, 11.28] |

### Association between ethnicity and the rate of disease progression

There was no consistent association pattern between ethnicity and disease accumulation, and this relationship was modified by comorbidity status (Figure 2). For CVD diagnosis across different comorbidity groups, (panel (a)), HRs tended to be slightly higher for South Asian but lower for Black when compared to White ethnicity. For T2D (panel (b)), HRs for South Asian, Black and Other ethnicities were significantly greater than 1 for most comorbidity groups, especially for the South Asian group (HRs ranging between 1.20 to 3.91), when compared to White. Furthermore, these HRs generally decreased with an increasing number of pre-existing conditions, as indicated by the fitted regression line from the meta-regression. In contrast, for CKD (panel (c)), HF (panel (d)) and MH (panel (e)) conditions, there were minor differences between the other ethnic groups and White across different transitions, with the HRs mostly close to 1. For MH, HRs for Black compared to White were lower than 1 across different comorbidity groups (HRs ranging between 0.52 to 0.92), and as observed for T2D, the differences between the two ethnic groups tended to decrease as the number of comorbidities increased.

**Figure 2.**
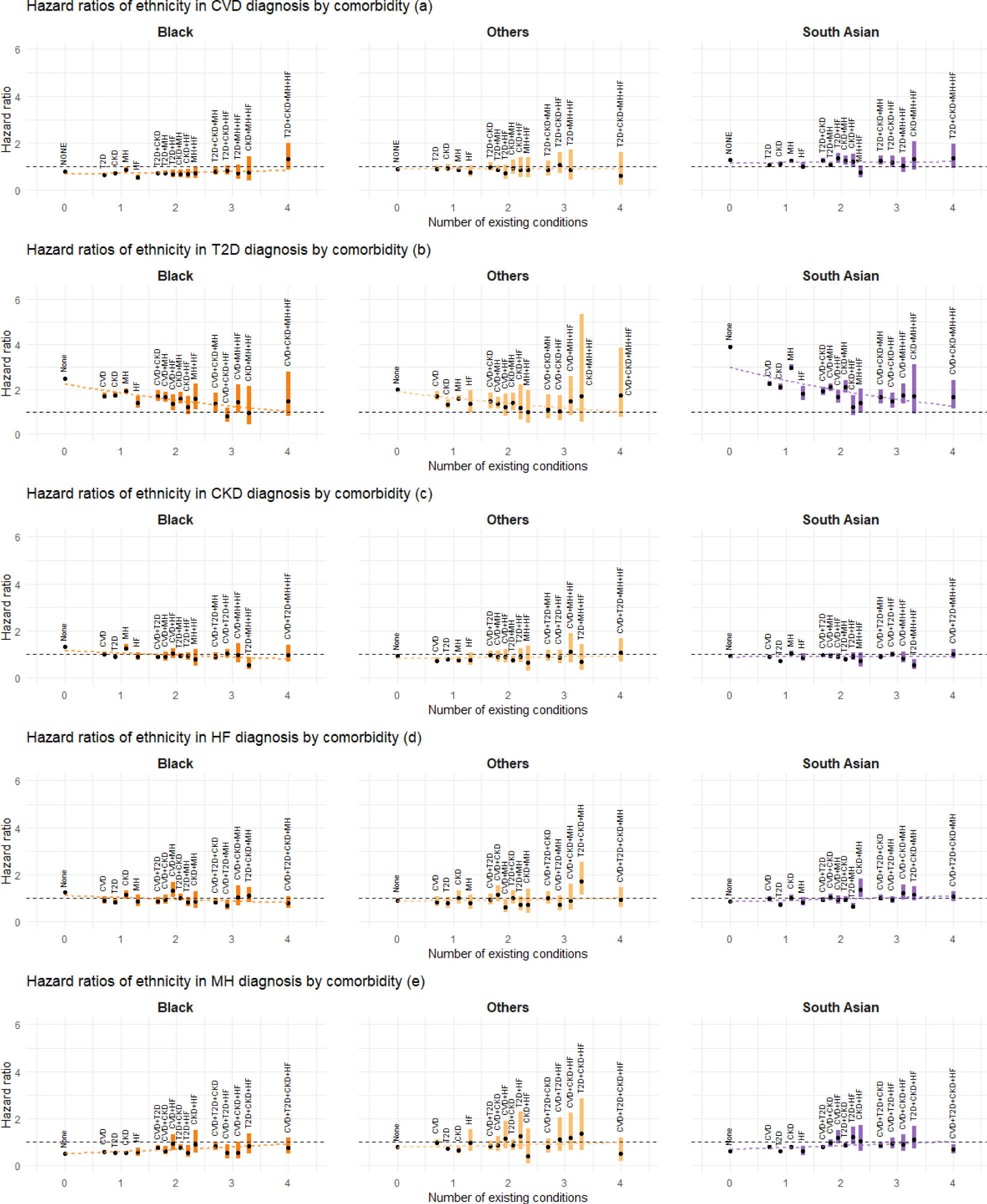
Estimated association between ethnicity (non-White groups) and the rate of disease transition by the number of existing conditions and comorbidity status. Estimated HRs are shown by black dots, with the associated 95% confidence intervals (CI) represented by coloured bands. White ethnicity is treated as the reference category. For transitions into each condition, the superimposed dotted lines represent meta-regression fits based on the corresponding HRs for each ethnicity category. Abbreviations: CVD: cardiovascular disease; T2D: type-2 diabetes; CKD: chronic kidney disease; HF: heart failure; MH: mental health conditions.

### Association between deprivation and the rate of disease progression

Across all types of disease progression, HRs for the more deprived quintiles (2nd to 5th IMD quintiles) compared to the least deprived quintile (1st IMD quintile) were mostly greater than 1. As the level of deprivation increased, the HRs typically increased, indicating a higher rate of disease transition (Figure 3). The magnitude of this association was generally larger for transitions into T2D (HRs for the 5th IMD quintile ranged between 0.95 to 2.03) and HF (HRs for the 5th IMD quintile ranged between 1.25 to 1.65), whereas it was weaker for CKD, MH, and CVD. The fitted meta-regression line further suggested that for CVD, T2D and MH, the association between greater deprivation and rate of disease accumulation tended to be attenuated with an increasing number of comorbidities. In contrast, this pattern was less evident for CKD and HF diagnosis.

**Figure 3.**
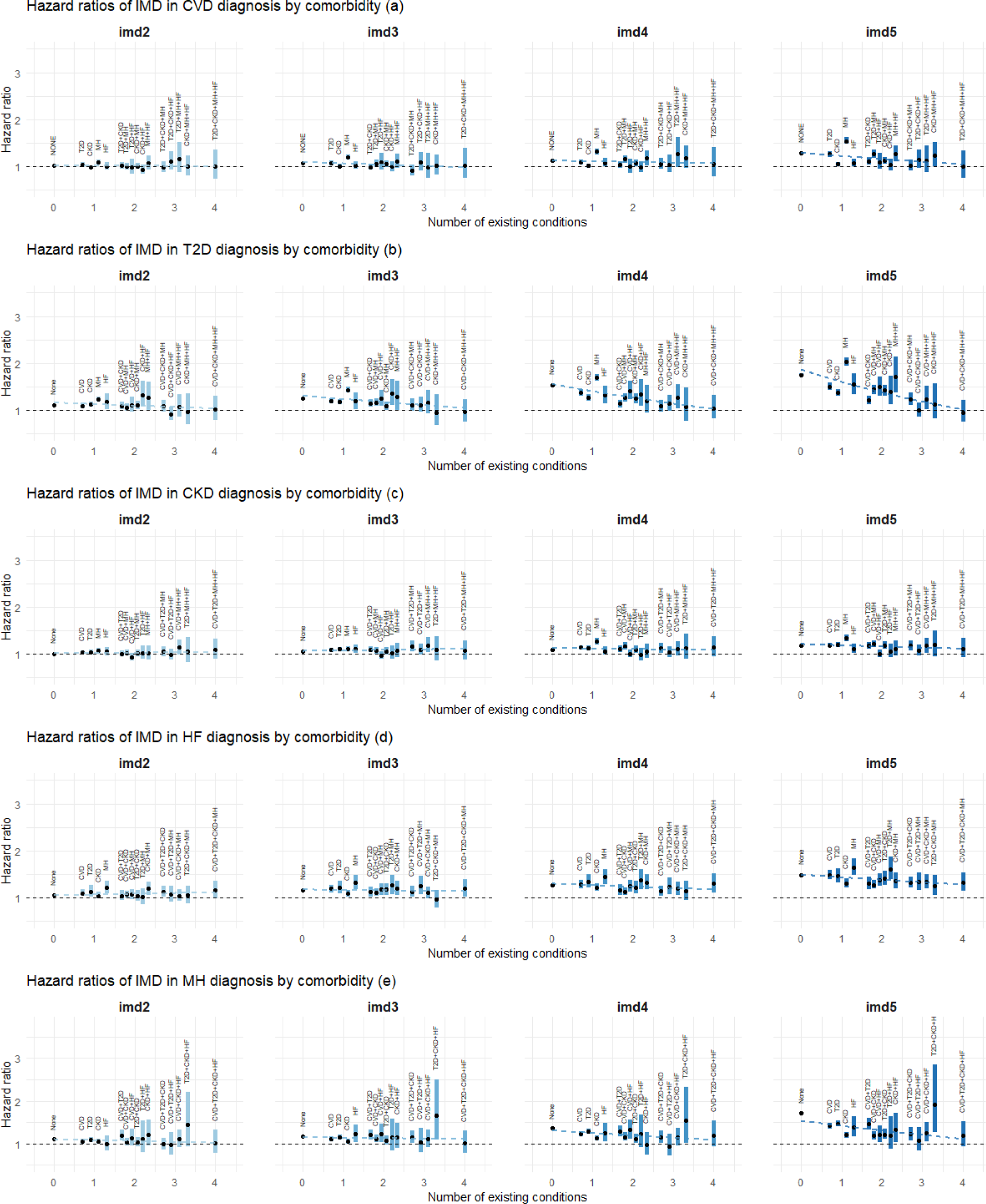
Estimated association between deprivation (2nd to 5th IMD quintiles) and the rate of disease transition by the number of existing conditions and comorbidity status. Estimated HRs are shown by black dots, with the associated 95% confidence intervals (CI) represented by coloured bands. The first IMD quintile (the least deprived group) is treated as the reference category. For transitions into each condition, the superimposed dotted lines represent meta-regression fits based on the corresponding HRs for each IMD category. Abbreviations: IMD: the English Index of Multiple Deprivation; CVD: cardiovascular disease; T2D: type-2 diabetes; CKD: chronic kidney disease; HF: heart failure; MH: mental health conditions; imd2 – imd5: 2nd - 5th IMD quintiles.

### Association between gender and the rate of disease progression

HRs for men, compared to women, were significantly higher than 1 for diagnoses of CVD (HRs ranging between 1.19 to 2.04), T2D (HRs ranging between 1.01 to 1.59) or HF (HRs ranging between 1.29 to 1.89), while they were lower than 1 for CKD (HRs ranging between 0.77 to 0.92) and MH (HRs ranging between 0.58 to 0.79) diagnoses across different comorbidity groups (Figure 4). The meta-regression analysis revealed that for CVD, T2D and HF diagnoses, the HRs for men exhibited a downward trend towards the null as the number of comorbidities increased, while for MH diagnosis, the initially lower HRs for men showed a slight upward adjustment with an increasing number of comorbidities. In contrast, for CKD diagnosis, there was little difference between men and women regardless of the number of pre-existing conditions.

**Figure 4.**
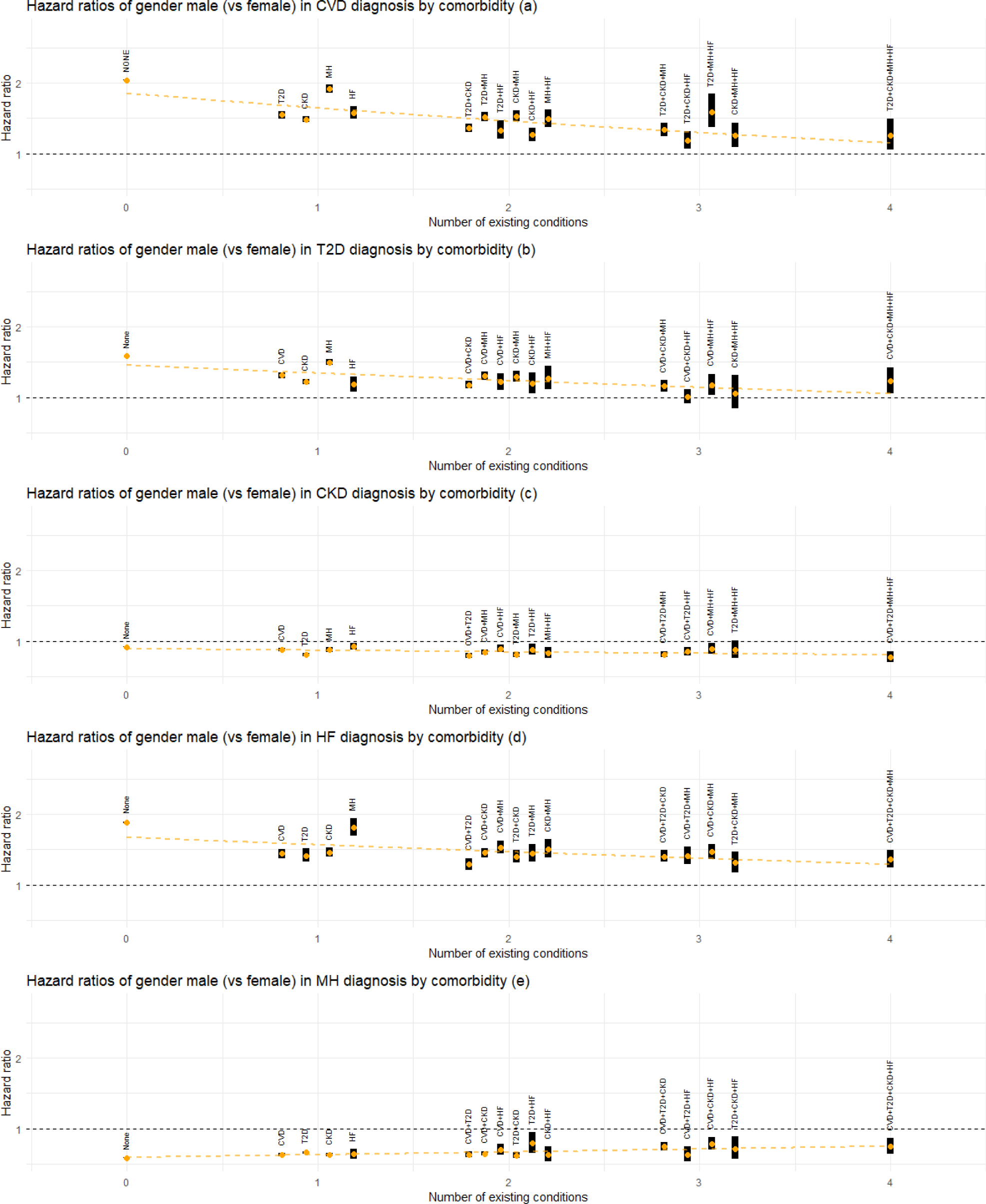
Estimated association between gender (male) and the rate of disease transition by the number of existing conditions and comorbidity status. Estimated HRs are shown by orange dots, with the associated 95% confidence intervals (CI) represented by black bands. The female gender is treated as the reference category. For transitions into each condition, the superimposed dotted line represents meta-regression fit based on the corresponding HRs. Abbreviations: CVD: cardiovascular disease; T2D: type-2 diabetes; CKD: chronic kidney disease; HF: heart failure; MH: mental health conditions.

### Association between age and the rate of disease progression

A 10-year increase in age was associated with an elevated rate of CVD (HRs ranging between 0.97 to 2.04), CKD (HRs ranging between 1.31 to 2.48), or HF (HRs ranging between 1.17 to 2.39) diagnosis (Figure 5). This increase was especially pronounced for patients with a single pre-existing condition, particularly MH, and tended to approach the null as the number of comorbidities increased. Conversely, HRs for a 10-year age increase were lower than 1 for MH diagnosis (HRs ranging between 0.75 to 0.92), and for T2D diagnosis except for the transition from MH (HRs ranging between 0.74 to 0.98). With the accumulation of comorbidities, the initially lower HRs for age showed a slight upward trend towards 1 for MH diagnosis, while an opposite trend was observed for transitions into T2D, where the HRs seemed to diverge further from 1 as the number of conditions increased.

**Figure 5.**
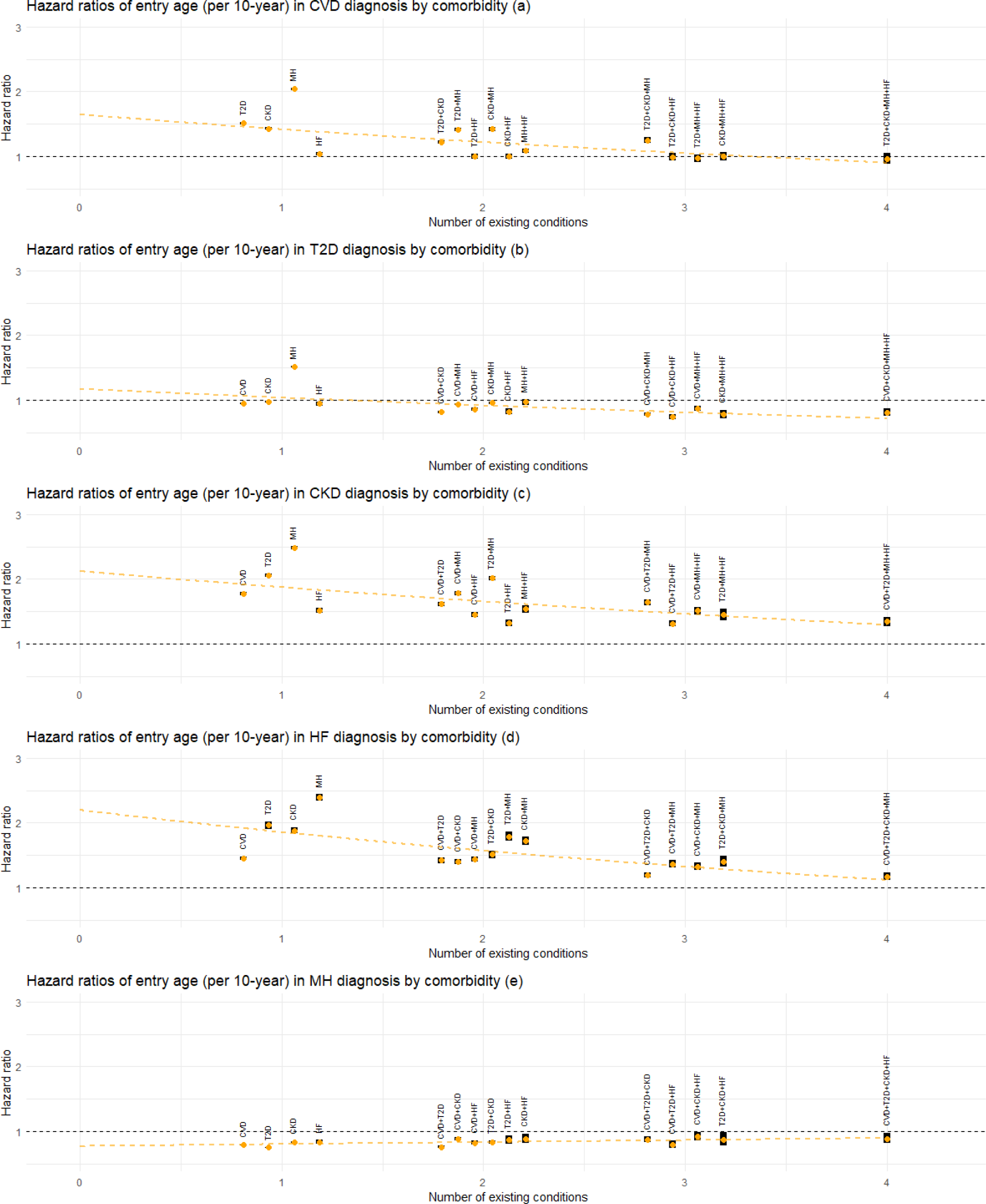
Estimated association between age at entry to the current state (per 10 years) and the rate of next disease transition by the number of existing conditions and comorbidity status. Estimated HRs are shown by orange dots, with the associated 95% confidence intervals (CI) represented by black bands. For transitions into each condition, the superimposed dotted line represents meta-regression fit based on the corresponding HRs. Abbreviations: CVD: cardiovascular disease; T2D: type-2 diabetes; CKD: chronic kidney disease; HF: heart failure; MH: mental health conditions.

### Mental health

Figures 2-5 consistently demonstrate a trend where the association between deprivation, gender, and age and the progression to multimorbidity tends to be more pronounced (in the direction of increasing the rate of transition) when transitions emerge from a mental health condition compared to other comorbidity states. For age and deprivation (particularly age), this pattern is observed for all transitions from MH. For instance, HRs (95% CI) for age were 2.04 (2.02, 2.06), 2.48 (2.45, 2.51) and 2.39 (2.34, 2.45) for transitions into CVD, CKD and HF, respectively. For transitions into T2D, the association with age becomes larger than 1 when the comorbidity status is MH (HR (95% CI): 1.52 (1.50, 1.53)), whereas it is less than 1 otherwise. The adverse association of being male (vs female) with transitions to CVD, T2D, or HF is amplified in cases involving a MH comorbidity, with HRs (95% CI) being 1.92 (1.86, 1.97), 1.50 (1.46, 1.54) and 1.82 (1.70, 1.95) respectively. For ethnicity, this pattern is only seen for transitions to T2D, where the HR (95% CI) for South Asian ethnicity (compared to White) for the transition from MH is 2.97 (2.86, 3.09).

### Sensitivity analysis results

Supplemental Figure C shows the HRs for each sociodemographic characteristic, focusing on transitions into CVD or MH based on the respective narrower definitions specified in the statistical methods section.

Modifying the conditions leads to some discrepancies in HRs, particularly for CVD. For instance, the gender (male) association for CVD diagnosis was stronger when focusing solely on IHD, while the age association was more pronounced for stroke/TIA. Despite this, the key patterns from our main results persisted, such as the attenuation of the HRs for deprivation, gender (male), and age with an increasing number of pre-existing conditions.

Results of the multistate analysis with COPD replacing HF are shown in Supplemental Figure D. Focusing on the disease transitions involving only CVD, T2D, CKD and MH conditions, we observed no noticeable differences from the results shown in Figures 2-5. Supplemental Figure E shows the results of the multistate analysis with COPD included as the sixth condition, focusing on the same transition types as in Figures 2-5. We see that the HRs are robust to the addition of a new condition in the transition network.

Supplemental Figure F shows the HRs of IMD deciles, and a comparison with Figure 3 (HRs of IMD quintiles) reveals very similar association patterns in more deprived groups. The larger CIs with the former is a result of the smaller sample size per decile category. The associations between ethnicity, age and gender, and disease progression remained essentially unaltered by using IMD deciles instead of quintiles. In fact, there is statistical evidence supporting the use of IMD quintiles rather than deciles, as the Akaike information criterion (AIC) favoured the former for the majority of the transitions.

Supplemental Figure G shows the HR results for the MSM fitted with an expanded cohort that includes those with missing ethnicity information (due to the computational time limitation of the HPC, we were not able to obtain HRs for transitions from ‘None’). The key patterns remained unchanged, and there were no substantial differences in the HRs when compared with Figures 2-5 in the main paper.

Supplemental Figure H shows the HR results for the MSM applied to our study cohort, specified based on semi-parametric Cox models. These results are substantively the same as those obtained under our spline-based parametric model (Figures 2-5).

## Discussion

### Main findings

In this study, we examined the associations between ethnicity, deprivation, gender, and age, and the rate of progression of five major chronic conditions, namely CVD, T2D, CKD, MH and HF, using a cohort of more than 13 million English adults. Our results revealed a complex interplay between these key sociodemographic characteristics and the progression of chronic diseases, with the strength and pattern of associations varying according to disease transition types. Deprivation, gender and age generally demonstrated stronger associations with the diagnosis of a condition than ethnic group differences. For some conditions, the impact of ethnicity, deprivation, gender, and age on the rate of transition attenuated with an increasing number of pre-existing conditions. For example, the rate of developing T2D is much higher in the most socially deprived groups and in those of South Asian and Black ethnicity compared to the reference groups, but the difference becomes smaller for those with four chronic conditions. This pattern also applies to the diagnosis of CVD or T2D in men, and the diagnosis of CVD, CKD or HF at younger ages. In addition, the impact of deprivation, gender, and age on the rate of diagnosis of CVD, T2D, CKD or HF was typically more pronounced when transitioning from a single mental health condition compared to a single physical health condition or other composite conditions. The robustness of our findings was confirmed through various sensitivity analyses.

### Contribution to the literature

Our findings align with and reinforce established epidemiological evidence. For instance, our observation of a higher rate of progression to T2D in patients of South Asian and Black ethnicities, compared with the White UK population [28]. Similarly, our findings regarding a higher rate of CVD progression in South Asian ethnicity and a lower rate in the Black population echo the patterns reported in the same study [28]. Our results show that higher levels of deprivation are generally associated with an elevated rate of disease progression across all conditions and transitions. This is consistent with previous research findings, which showed that multimorbidity occurred 10–15 years earlier and physical-mental comorbidity was over twice as prevalent in the most socioeconomically deprived areas of Scotland than in the least deprived areas [18]. With regard to gender, our results align with previous studies, indicating men generally have an increased risk of progression to CVD, T2D, and HF, but exhibit a lower risk of MH problems and CKD [20]. Moreover, our study further corroborates well-established findings that ageing is strongly associated with an increased rate of CVD, CKD and HF [29].

Our multistate analysis offers novel insights by revealing a detailed association between sociodemographic characteristics and disease progression (CVD, T2D, CKD, MH and HF), thereby underlining a complex dependency on the underlying comorbidity status prior to the transition. In particular, we found that the association between these characteristics and disease transitions tended to be greater when the number of pre-existing conditions was lower. This implies that the impact of these characteristics may be larger during initial multimorbidity development and decrease in later stages. This pattern was particularly noticeable for ethnicity (for transitions into T2D), deprivation (for transitions into CVD, T2D and MH), age (for transitions into CVD, CKD, MH and HF), and gender (for transitions into CVD, T2D, MH and HF). With regard to ethnicity and deprivation, T2D stood out from the other four conditions due to a stronger influence/attenuation of these factors by the number of pre-existing conditions on T2D diagnosis. A plausible explanation for this pattern could be that as patients accumulate more conditions, the cumulative effect of these existing conditions may become the predominant factor influencing further disease accumulation. Additionally, enhanced health management and monitoring, which may include lifestyle modifications and medical interventions/treatments, could counterbalance the impact of sociodemographic factors.

Another novel finding is that the adverse impact of sociodemographic factors such as deprivation, gender, and age on the rate of transitioning to the next condition (CVD, T2D, CKD or HF) tended to be more pronounced when transitioning from MH alone compared to other single or composite conditions. This pattern may again be partly attributed to biological and behavioural factors. Patients with MH as their starting condition tend to fall into the younger age groups (see Supplemental Table A) and mental ill health is associated with unhealthy lifestyle habits such as smoking, alcohol or substance abuse among others [30], which are known to be important risk factors for cardiometabolic diseases and contribute to the worsening of multimorbidity [13,31]. On the other hand, this adverse influence of MH appears to diminish with the presence of additional comorbidities, which could be due to similar reasons mentioned earlier.

### Strengths and limitations

This study benefits from a large dataset from primary health care that is broadly representative of the English population. The five chronic conditions considered in this study are included in an incentive scheme for the management of chronic conditions, which means they are likely to be more accurately coded [32]. We employed a flexible multistate model to comprehensively examine the association between key sociodemographic characteristics and the rate of different disease transitions during multimorbidity accumulation involving the five specified conditions. The model achieved a good overall fit for the vast majority of the transitions, demonstrating the utility and computational feasibility of the approach. To our knowledge, this is the largest cohort and the highest number of chronic conditions and disease transitions analysed using a flexible multistate modelling framework.

While our primary analysis centers on five specific conditions, our modelling and estimation approach can be adapted to accommodate different or additional conditions. The complexity of the corresponding MSM (under the same assumptions on the permitted direct transitions as described in the Methods section) grows exponentially with N, the total number of conditions under consideration. However, this rise in complexity does not imply a proportional increase in computational costs. By leveraging a valid parallel computing strategy, we estimated each transition independently, and observed that for the same dataset, including more conditions typically leads to fewer observed events per transition (due to the increase in competing events for each transition). In our sensitivity analysis, we were able to fit a MSM that included a sixth condition, COPD, within our computational budget and importantly, the results of interest remained robust with the inclusion of COPD. We anticipate similar outcomes with the inclusion of other conditions. Nevertheless, depending on the scope of transitions of interest, a more complex MSM could pose challenges in summarizing, presenting, and interpreting results. On the other hand, our sensitivity analysis indicates that the estimated MSM could be influenced to some extent by modifications to the condition definition. This is expected, given that we define the diagnosis time of a composite condition (such as CVD and MH) as the first occurrence of any associated sub-conditions. Therefore, any change in the set of sub-conditions could have a direct and complex impact on a patient’s event history data. In our sensitivity analysis, we observed greater differences in results when changing the definition of CVD compared to MH. This is partly due to the smaller overlap between CVD subtypes (stroke/TIA and IHD) as opposed to MH subtypes (anxiety and depression). In addition, we observed a closer alignment of results from the main analysis when using CVD (IHD only) or MH (depression only) definitions. This is because in our dataset, the presence of IHD and depression contributed to a larger proportion of the diagnosis of CVD and MH (using our original definitions), respectively, compared to the alternative definitions. While there are no universally accepted definitions for complex chronic conditions like CVD or MH, we believe that their determination should be considered in the context of the research question of interest and the data available.

It should be pointed out, however, that potential diagnostic inaccuracies in primary care records, such as underdiagnosis, overdiagnosis, or ascertainment bias of chronic conditions, could influence our results. For instance, diseases like CKD and T2D are often asymptomatic in the early stages, and diagnosis may occur during routine health check/screening or following the diagnosis of a related condition [33,34]. It is noted that the uptake of such checkups is generally less than 50%, with gender and ethnic disparities in the presentation to healthcare - specifically, lower uptake among men and some ethnic groups [35,36]. For instance, for CKD, renal function testing is less common in minority ethnic groups [37], and for MH, certain groups (e.g. those in the Black or South Asian ethnic groups) are less likely to present to the GP even when symptoms have developed [38]. The influence of these potential inaccuracies might be more pronounced during the early phases of multimorbidity, as participants with existing health conditions could be more likely to be invited for (and perhaps more inclined to undergo) further monitoring and screening for other health conditions. We note that for MH in particular, studies have reported significant challenges with both under and over-reporting of depression and anxiety in primary care records. Other studies have used algorithms involving prescription data, symptom and diagnosis clinical codes to improve case detection of MH conditions [18,39,40], whereas we only used diagnosis codes. A preprint study using the same coding strategy in this same database has compared the prevalence of depression in the CPRD Aurum database with the prevalence reported in a representative sample of the English population in the Adult Psychiatric Morbidity Survey (APMS) [20,41]. Prevalence of depression in >16 year olds in CPRD Aurum was similar (19.5%; 95% CI: 19.5 - 19.5) to the doctor diagnosed depression prevalence in APMS (20.9%; 95% CI: 20.0 - 21.8) but lower than the prevalence of symptoms of depression (27.8%; 95% CI: 26.9 - 28.8). On the other hand, we acknowledge that participants who are less likely to present to primary care or receive diagnostic labels, such as socially deprived younger males, may have absent MH data. This could lead to an underrepresentation of participants from deprived communities. If anything, this may suggest that our findings – such as the association between deprivation and the rate of transition to the next condition is typically more pronounced when transitioning from a mental health condition - may actually be understated.

There are some other limitations with this study. For instance, the ethnicity categories used in this study crudely group together communities with diverse cultural practices and sociodemographic characteristics, which may mask differences in outcomes within each category. Note also that time-varying factors associated with the disease progression, such as treatment information, lifestyle modifications, and change in deprivation status, are not available from the present dataset and thus they were not taken into account in the present analysis. Finally, despite our large cohort which provides a reasonable amount of data for estimating each transition, we observe a substantial decrease in the number of transitions as more conditions accumulate. This trend is reflected in the widening confidence interval bands in Figures 2-5, and is particularly noticeable with categorical variables like ethnicity, where an imbalance between groups exists. Consequently, this adds some level of uncertainty to our interpretations. For multimorbidity studies using a multistate model like ours, longer follow-up periods and/or larger datasets would always be desirable.

### Implications for policy and practice

The higher rates of development of the first chronic conditions in a number of identifiably disadvantaged population groups has two implications. First, it suggests that the factors contributing towards differences in the development of chronic conditions precede engagement with health services. This points towards wider determinants of health, particularly in relation to T2D and CVD. A better understanding of the differential impacts of socioeconomic factors on the development of chronic conditions would be valuable. Second, it implies that the greatest potential to reduce inequalities in multimorbidity between more and less socially deprived groups and between those of white and South Asian or Black ethnicities is early in the disease trajectory. Prevention of CVD and T2D might be an appropriate focus. Initiatives such as the Marmot cities are consistent with both these implications [42]. Evaluation of interventions to prevent the development of the first and second chronic conditions would be valuable. As multimorbidity, in particular mixed physical and mental health conditions, increases the use of secondary care services, an understanding of the economic effects of multimorbidity could help identify prevention priorities [42–44].

### Future directions

Clinical research implications of this analysis are to investigate and better understand the biopsychosocial factors driving the progression to multimorbidity. The greatest differences in transition rates to multimorbidity between different groups are in the development of the first few conditions, which suggests that the early stages of multimorbidity development could be particularly important and require increased attention.

Future work could apply this method to analysing the trajectories of different clusters of diseases. These might include other common conditions such as cancer, asthma and hypertension or rarer, less explored conditions. It could also investigate groups of potentially related conditions such as mental health problems, autoimmune conditions or medically unexplained symptoms. This would help broaden the scope of multimorbidity research. From a methodological perspective, the MSM framework we utilised can be extended to address additional questions pertinent to longitudinal multimorbidity research. Ongoing work is developing a scalable and flexible joint modelling framework for both the longitudinal and multistate processes [45]. This will facilitate the investigation of associations between concurrently measured longitudinal covariates (such as biomarkers) and the progression of multimorbidity. Another direction involves the integration of modern deep learning techniques into MSM to enhance its flexibility and predictive accuracy for micro-simulation. This approach will enable the generation and analysis of longitudinal multimorbidity trajectories at both individual and population levels.

## Conclusion

Multimorbidity is a growing public health challenge and there is a need to shift from a single disease-oriented approach to a more individualised and holistic strategy. In this study, we examined the association between key sociodemographic characteristics and the progression of multimorbidity involving five common chronic conditions: namely CVD, T2D, CKD, HF and MH. Our findings reveal distinctive association patterns across different disease transitions. Notably, the impact of sociodemographic factors tends to attenuate with an increasing number of pre-existing conditions, with patterns varying depending on the specific diagnosed condition. Mental health, as a comorbidity, showed unique patterns, with the impact of deprivation, gender, and age typically more pronounced when transitioning from a mental health condition. These findings underscore the potential importance of earlier detection and intervention of chronic diseases, especially mental health conditions, and particularly for those in the higher-risk populations. These insights may have important implications for improved patient care, socioeconomic considerations, and healthcare strategies.

## Data Availability

The data were provided under license by the Clinical Practice Research Datalink (CPRD), and so are not publicly available. Access to CPRD data is subject to protocol approval via CPRD's Research Data Governance (RDG) Process.

## Acknowledgements

This work used data from the Clinical Practice Research Datalink (CPRD) for ISAC protocol 19_265. The data were provided by patients and collected by the NHS as part of their care and support.

For the purpose of open access, the author has applied a Creative Commons Attribution (CC BY) licence to any Author Accepted Manuscript version arising from this submission.

## Author contributions

### Conceptualization

Tom Marshall, Francesca Crowe, Krish Nirantharakumar, Duncan Edwards, Jessica K Barrett

### Data curation

Catherine L Saunders Formal analysis: Sida Chen

### Funding acquisition

Tom Marshall, Francesca Crowe, Krish Nirantharakumar, Paul Kirk, Sylvia Richardson, Duncan Edwards, Simon Griffin, Christopher Yau, Jessica K Barrett

### Methodology

Sida Chen, Tom Marshall, Christopher Jackson, Jessica K Barrett Project administration: Sida Chen, Francesca Crowe

### Supervision

Jessica K Barrett

### Writing – original draft

Sida Chen, Tom Marshall, Jennifer Cooper

### Writing – review & editing

Sida Chen, Tom Marshall, Christopher Jackson, Jennifer Cooper, Francesca Crowe, Krish Nirantharakumar, Catherine L Saunders, Paul Kirk, Sylvia Richardson, Duncan Edwards, Simon Griffin, Christopher Yau, Jessica K Barrett

## Supplementary information legends

**S1 File. Supplementary material to the main manuscript (DOCX)**

Fig A. Flowchart for sample selection. Text. Supplemental Technical Details

Table A. Summary of disease states by sociodemographic characteristics. Table B. Summary of disease transitions.

Fig B. Plots of the estimated cumulative hazard of the Cox-Snell residuals.

Fig C. Estimated association between each sociodemographic characteristic and the rate of CVD or MH diagnosis based on their respective narrower definitions.

Fig D. Results of the multistate analysis obtained with COPD replacing HF.

Fig E. Results of the multistate analysis with COPD included as the sixth condition.

Fig F. Estimated association between deprivation (IMD deciles) and the rate of disease transition by number of existing conditions and comorbidity status.

Fig G. Results of the multistate analysis obtained based on the expanded cohort including patients with missing ethnicity information.

Fig H. Results of the multistate analysis with transitions specified via semi-parametric Cox models.

CVD: cardiovascular disease; HF: heart failure; MH: mental health conditions; COPD: chronic obstructive pulmonary disease; IMD: the English Index of Multiple Deprivation.

**S1 Code lists. Code lists for ethnicity and the conditions considered in our study. (ZIP)**

