## supplemental materials for "Sociodemographic Characteristics and Longitudinal Progression of Multimorbidity: A Multistate Modelling Analysis of a Large Primary Care Records Dataset in England"

#### Supplemental Figure A. Flowchart for sample selection

**Extraction stage from CPRD Aurum**

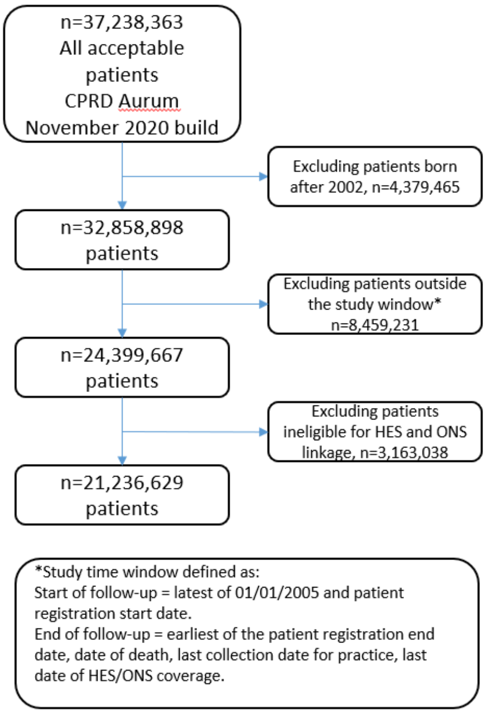


**Additional criteria applied at the data processing stage**

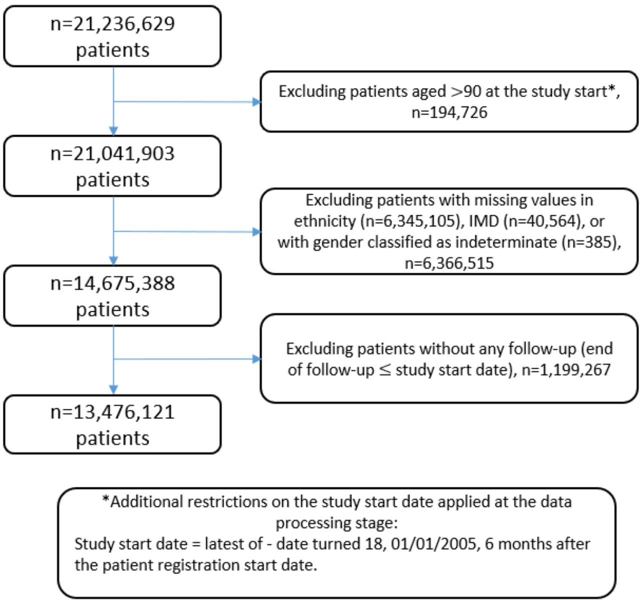

### Supplemental Technical Details

**The multistate modelling approach**

In this section we provide a brief introduction to multistate modelling (MSM), and we refer to [1] and references therein for more in-depth coverage of the topic. A multistate model is built from a mathematical object named a multistate process. A multistate process is a stochastic process $\{X(t),\ t \geq 0\}$ with a finite state space $S = \{1, \ldots, N\}$ (here we focus on the setting where $t$ is continuous and $X(t)$ is continuously observed). The law of a multistate process can be fully characterised via the transition intensities, which directly describe the instantaneous dynamic of the process

$$\alpha_{hj}(t; F_{t^-}) = \lim_{\Delta t \to 0} \frac{P(X(t + \Delta t) = j | X(t) = h;\ F_{t^-})}{\Delta t}\ ,\ \ h \neq j \in S,$$

where $F_{t^-}$ represents the history of the process over $[0, t)$. There are two special classes of the multistate processes that have been thoroughly studied. The first is the Markov processes, which relies on the Markov assumption such that risks at a particular time are independent of the history, so that

$$\alpha_{hj}(t; F_{t^-}) = \alpha_{hj}(t).$$

The second class is the so-called clock-reset semi-Markov processes, which relaxes the Markov property by assuming that

$$\alpha_{hj}(t; F_{t^-}) = \alpha_{hj}\big(B(t)\big),$$

where $B(t)$ is the time since entry to the current state h. Therefore, $\alpha_{hj}$ depends on the history only through $B(t)$, and in effect, the time scale after each transition is reset to zero. In the former scenario, the transition probabilities between two states can be derived from the intensities by solving the Kolmogorov differential equations, whereas for the latter case, no explicit formulation exists. In such scenarios, one could estimate transition probabilities from intensities via simulation-based techniques.

A multistate model can be fitted to data representing the evolution of $n$ subjects (defined on the same state space) in a population, each of whom is followed up continuously for a potentially different length of time (may be subject to delayed entry or right censoring). To model the covariate-dependent transition intensities for subject $i$, $\alpha_{hj}^{(i)}(t; F_{t^-})$, Markov or semi-Markov assumptions are commonly adopted and semi-parametric or parametric proportional hazard regression framework is often used. If we in addition assume that each individual's trajectory, $X_i(t)$, is independent, then it can be shown using counting process theory that the MSM likelihood factorizes as

$$L = \prod_{i=1}^{K} L_i\big(\theta | D^{(i)}\big),$$

where $K$ is the total number of allowed transition types in the multistate process (e.g. if $\alpha_{hj} \neq 0$ then $h \rightarrow j$ represents one type of transition), $L_i$ is the likelihood obtained by considering transition type $i$ as a survival problem with data based on all subjects at risk for this transition and $\theta$ is the parameter vector consisting of parameters from all transitions. An important consequence of this factorization property is that if no parameters are shared across transitions, i.e. $\theta = (\theta_1, \ldots, \theta_K)$, then we could perform maximum likelihood estimation of the MSM by maximising each $L_i$ independently with respect to its parameter set $\theta_i$ (see e.g. [2]).

In our analysis, we specify the MSM by using a separate time-to-event model for each permitted $h \to j$ transition. To ensure flexibility, we use the spline-based Royston-Parmar model [3] for the transition-specific intensities, $\alpha_{hj}^{(i)}(u; Z(t))$,

$$log\ (A_{hj}^{(i)}(u; Z)) = s_{hj}(log\ u\ ; \gamma_{hj}) + \beta_{hj}^{T} Z_i\ ,$$

where $u$ refers to the time since entry to the state $h$, $A_{hj}^{(i)}$ is the cumulative transition intensity associated with $\alpha_{hj}^{(i)}$, $s_{hj}(log\ u\ ; \gamma_{hj})$ is a natural cubic spline in $log\ u$ with coefficient vector $\gamma_{hj}$ (see [3] for more details on the construction of the spline function), $Z_i$ denotes the vector of the exogenous covariates for subject $i$ and $\beta_{hj}$ is the vector of the associated linear effects. Note that the resulting time-to-event model retains a proportional hazard structure so that elements of $\beta_{hj}$ are still interpreted as log-hazard ratios. In our context, the covariates/sociodemographic characteristics are IMD, ethnicity, gender and age (at the time of entering the current state; in 10 years). The first three variables are categorical and are represented via dummy variables with reference groups set to female, the first IMD quintile (the least deprived group) and White (the dominant ethnic group), respectively. Age is approximated by a piecewise constant process with jumps occurring at the transition times (hence is treated as a time constant covariate in a transition model). A practical issue here is to determine the number and location of the internal knots for the spline term $s_{hj}$, which would influence the flexibility of the resulting model. We follow the suggestions in [3] to use the Akaike information criterion (AIC) to determine the number of internal knots in the spline from the candidate values of $\{0, \ldots, 5\}$. Note that when there are zero knots the model reduces to a Weibull model. Conditional on the knot number, the location of these knots is set to equally spaced percentiles of the logarithm of the uncensored transition times for $h \to j$ (empirical study indicates that the knot position is not a key driver of model fit).

**Model checking**

We used the Cox-Snell residuals to graphically examine the goodness of fit of each of our transition models [4,5]. For a fitted time-to-event model, the Cox-Snell residuals are defined as $r_i^{CS} = \hat{A}_i(T_i)$, where $\hat{A}_i$ is the estimated cumulative hazard function for subject $i$ under the fitted model and $T_i$ is the observed event time. If the model fits the data well, we expect the $r_i^{CS}$ to be close to draws from a unit exponential distribution. This diagnostic is motivated by the fact that for any random variable $T$ with survival function $S(t) = P(T > t)$, the transformed random variable $-\ log\ S(T)$ is distributed as an exponential random variable with unit mean. To appropriately take censored event times into account, in practice we estimate the cumulative hazard of the residuals, i.e. $-\ log\ \hat{S}\ (r_i^{CS})$, where $\hat{S}$ is the Kaplan-Meier estimate of the survival function associated with $r_i^{CS}$ (for censored event times $T_i$ the corresponding $r_i^{CS}$ are treated as censored as well). A good fit is suggested if a plot of $-\ log\ \hat{S}\ (r_i^{CS})$ against $r_i^{CS}$ is close to a straight line with an intercept of zero and a slope of one. For our fitted MSM, we computed the Cox-Snell residuals for each transition, and we observed satisfactory results for most of the transitions, see Figure B for selective results. Here we observed a slight lack of fit for transitions MH->MH+T2 and MH->MH+CK. We found that residuals that significantly deviated from the reference line (in red) correspond to patients who had a long transition time, but their sociodemographic characteristics suggest that the transition should have happened earlier. The presence of such “outliers” may not be surprising given the complexity of the disease progression mechanisms and the inter-subject heterogeneity in such a large cohort. On the other hand, it should be noted that these outliers only account for an extremely small proportion of the data involved in these 112 transitions. Out of the 112 transitions these two were among the most ill-fitting.

**Meta-regression**

For a given sociodemographic characteristic and for transitions into diagnosis of a specific condition, we considered the following meta-regression model:

$$log\,\widehat{HR}_i = \beta_0 + \beta_1 level_i + v_i + \epsilon_i,$$

where $\widehat{HR}_i$ is the estimated hazard ratio for a specific group of the characteristic associated with transition type i, $level_i$ denotes the count of pre-existing conditions prior to transition type i, $v_i \sim N(0, \tau^2)$ is the transition-specific random effect, with $\tau^2$ being the between-transition variance, and $\epsilon_i \sim N(0, \hat{\sigma}_i{}^2)$ represents the within-transition random error, where $\hat{\sigma}_i$ is the estimated standard error associated with $\widehat{HR}_i$. Within our setting, $level_i$ can have values ranging from 0 to 4 (as we have a total of five conditions). We did not consider more complex models as there are only 16 HRs for each category of the characteristics, each HR corresponds to a transition associated with a diagnosis of a specific condition (note that for age, there are only 15 HRs linked to it as when $level_i = 0$, age is zero). In our implementation, we used the metafor R package. The model parameters were estimated via the Restricted Maximum Likelihood (REML) method, and the Knapp-Hartung method was used for adjusting the standard errors of the estimated effect sizes to provide more conservative estimates. The intercept ($\beta_0$) and slope ($\beta_1$) parameters together characterize the potential impact of the number of conditions on the hazard ratios. The fitted regression line, transformed back to the original hazard ratio scale via $exp\,(\hat{\beta}_0 + \hat{\beta}_1 level_i)$, is superimposed on our hazard ratio plots (e.g. Figures 2-5) to facilitate illustrating the relationship.

Supplemental Table A. Summary of disease states by sociodemographic characteristics.

| | | age* | IMD | | | | | Gender | | Ethnicity | | | | |
|---|---|---|---|---|---|---|---|---|---|---|---|---|---|---|
| Disease state | Counts | Median (IQR) | 1st Quintile (Least deprived) | 2nd Quintile | 3rd Quintile | 4th Quintile | 5th Quintile | Male | Female | White | Asian | Black | Mixed | Others |
| CV | 207102 | 68.4 [58.7, 77.2] | 45889 (22.2%) | 44310 (21.4%) | 41403 (20%) | 38687 (18.7%) | 36813 (17.8%) | 128239 (61.9%) | 78863 (38.1%) | 189530 (91.5%) | 10563 (5.1%) | 4781 (2.3%) | 1194 (0.6%) | 1034 (0.5%) |
| T2 | 269791 | 58.4 [48.7, 68.3] | 44150 (16.4%) | 47236 (17.5%) | 51680 (19.2%) | 62833 (23.3%) | 63892 (23.7%) | 158576 (58.8%) | 111215 (41.2%) | 196363 (72.8%) | 44772 (16.6%) | 21457 (8%) | 4157 (1.5%) | 3042 (1.1%) |
| CK | 211219 | 74.3 [65.9, 81.3] | 47907 (22.7%) | 45631 (21.6%) | 43041 (20.4%) | 39080 (18.5%) | 35560 (16.8%) | 85293 (40.4%) | 125926 (59.6%) | 195653 (92.6%) | 6810 (3.2%) | 6808 (3.2%) | 1333 (0.6%) | 615 (0.3%) |
| MH | 947978 | 37.3 [27.3, 49.8] | 166803 (17.6%) | 177562 (18.7%) | 179870 (19%) | 205494 (21.7%) | 218249 (23%) | 347552 (36.7%) | 600426 (63.3%) | 835101 (88.1%) | 56324 (5.9%) | 31355 (3.3%) | 14755 (1.6%) | 10443 (1.1%) |
| HF | 35434 | 73.7 [62.9, 82.1] | 7283 (20.6%) | 7195 (20.3%) | 7173 (20.2%) | 6960 (19.6%) | 6823 (19.3%) | 20587 (58.1%) | 14847 (41.9%) | 32670 (92.2%) | 1175 (3.3%) | 1221 (3.4%) | 221 (0.6%) | 147 (0.4%) |
| CV,T2 | 74537 | 69.7 [61.3, 77.3] | 12870 (17.3%) | 13932 (18.7%) | 14573 (19.6%) | 16050 (21.5%) | 17112 (23%) | 50521 (67.8%) | 24016 (32.2%) | 61380 (82.3%) | 8869 (11.9%) | 3029 (4.1%) | 753 (1%) | 506 (0.7%) |

| | | | | | | | | | | | | | |
|---|---|---|---|---|---|---|---|---|---|---|---|---|---|
| CV,CK | 93647 | 79.1 [72.7, 84.6] | 20775 (22.2%) | 20080 (21.4%) | 19178 (20.5%) | 17327 (18.5%) | 16287 (17.4%) | 48526 (51.8%) | 45121 (48.2%) | 89170 (95.2%) | 2543 (2.7%) | 1381 (1.5%) | 359 (0.4%) | 194 (0.2%) |
| CV,MH | 108335 | 65.8 [56.1, 75.6] | 19829 (18.3%) | 20968 (19.4%) | 21023 (19.4%) | 21996 (20.3%) | 24519 (22.6%) | 51951 (48%) | 56384 (52%) | 101998 (94.2%) | 3701 (3.4%) | 1558 (1.4%) | 584 (0.5%) | 494 (0.5%) |
| CV,HF | 29851 | 76 [67.2, 83] | 5940 (19.9%) | 6094 (20.4%) | 5962 (20%) | 5807 (19.5%) | 6048 (20.3%) | 20458 (68.5%) | 9393 (31.5%) | 27995 (93.8%) | 1107 (3.7%) | 521 (1.7%) | 116 (0.4%) | 112 (0.4%) |
| T2,CK | 73307 | 74.5 [67.3, 80.6] | 13278 (18.1%) | 14230 (19.4%) | 14731 (20.1%) | 15539 (21.2%) | 15529 (21.2%) | 34860 (47.6%) | 38447 (52.4%) | 61431 (83.8%) | 6039 (8.2%) | 4664 (6.4%) | 839 (1.1%) | 334 (0.5%) |
| T2,MH | 131774 | 56.9 [48.2, 66.2] | 18727 (14.2%) | 21816 (16.6%) | 24835 (18.8%) | 30130 (22.9%) | 36266 (27.5%) | 58203 (44.2%) | 73571 (55.8%) | 109628 (83.2%) | 13163 (10%) | 6137 (4.7%) | 1622 (1.2%) | 1224 (0.9%) |
| T2,HF | 11159 | 73 [63.9, 80.6] | 1781 (16%) | 2002 (17.9%) | 2228 (20%) | 2519 (22.6%) | 2629 (23.6%) | 6943 (62.2%) | 4216 (37.8%) | 9331 (83.6%) | 946 (8.5%) | 698 (6.3%) | 133 (1.2%) | 51 (0.5%) |
| CK,MH | 82217 | 71.8 [62.5, 80] | 17186 (20.9%) | 17042 (20.7%) | 16515 (20.1%) | 15764 (19.2%) | 15710 (19.1%) | 20981 (25.5%) | 61236 (74.5%) | 78644 (95.7%) | 1616 (2%) | 1391 (1.7%) | 390 (0.5%) | 176 (0.2%) |
| CK,HF | 17020 | 81.8 [74.9, 87] | 3581 (21%) | 3600 (21.2%) | 3540 (20.8%) | 3282 (19.3%) | 3017 (17.7%) | 7944 (46.7%) | 9076 (53.3%) | 16066 (94.4%) | 409 (2.4%) | 423 (2.5%) | 80 (0.5%) | 42 (0.2%) |
| MH,HF | 14443 | 70.1 [58.9, 79.8] | 2530 (17.5%) | 2752 (19.1%) | 2849 (19.7%) | 2995 (20.7%) | 3317 (23%) | 6339 (43.9%) | 8104 (56.1%) | 13762 (95.3%) | 288 (2%) | 274 (1.9%) | 62 (0.4%) | 57 (0.4%) |

| | | | | | | | | | | | | | | |
|---|---|---|---|---|---|---|---|---|---|---|---|---|---|---|
| CV,T2,CK | 42772 | 77.4 [71.4, 82.9] | 7618 (17.8%) | 8224 (19.2%) | 8537 (20%) | 8897 (20.8%) | 9496 (22.2%) | 24674 (57.7%) | 18098 (42.3%) | 36887 (86.2%) | 3713 (8.7%) | 1603 (3.7%) | 385 (0.9%) | 184 (0.4%) |
| CV,T2,MH | 39093 | 66.9 [58.6, 75.1] | 5544 (14.2%) | 6609 (16.9%) | 7195 (18.4%) | 8828 (22.6%) | 10917 (27.9%) | 21282 (54.4%) | 17811 (45.6%) | 34012 (87%) | 3393 (8.7%) | 1102 (2.8%) | 326 (0.8%) | 260 (0.7%) |
| CV,T2,HF | 13867 | 74.3 [66.8, 80.8] | 2135 (15.4%) | 2452 (17.7%) | 2700 (19.5%) | 3099 (22.3%) | 3481 (25.1%) | 10050 (72.5%) | 3817 (27.5%) | 11865 (85.6%) | 1339 (9.7%) | 489 (3.5%) | 106 (0.8%) | 68 (0.5%) |
| CV,CK,MH | 38653 | 78.1 [70.8, 84.3] | 7634 (19.8%) | 7799 (20.2%) | 7689 (19.9%) | 7577 (19.6%) | 7954 (20.6%) | 13949 (36.1%) | 24704 (63.9%) | 37386 (96.7%) | 721 (1.9%) | 350 (0.9%) | 128 (0.3%) | 68 (0.2%) |
| CV,CK,HF | 23222 | 81.5 [75.6, 86.6] | 4713 (20.3%) | 4788 (20.6%) | 4765 (20.5%) | 4438 (19.1%) | 4518 (19.5%) | 13777 (59.3%) | 9445 (40.7%) | 22115 (95.2%) | 631 (2.7%) | 335 (1.4%) | 90 (0.4%) | 51 (0.2%) |
| CV,MH,HF | 13155 | 73.8 [64.1, 81.6] | 2166 (16.5%) | 2440 (18.5%) | 2587 (19.7%) | 2745 (20.9%) | 3217 (24.5%) | 7389 (56.2%) | 5766 (43.8%) | 12532 (95.3%) | 358 (2.7%) | 168 (1.3%) | 54 (0.4%) | 43 (0.3%) |
| T2,CK,MH | 28205 | 72.4 [64.6, 79.5] | 4703 (16.7%) | 5280 (18.7%) | 5437 (19.3%) | 5951 (21.1%) | 6834 (24.2%) | 9266 (32.9%) | 18939 (67.1%) | 25225 (89.4%) | 1550 (5.5%) | 1083 (3.8%) | 224 (0.8%) | 123 (0.4%) |
| T2,CK,HF | 8441 | 79.1 [72, 84.7] | 1373 (16.3%) | 1586 (18.8%) | 1741 (20.6%) | 1828 (21.7%) | 1913 (22.7%) | 4491 (53.2%) | 3950 (46.8%) | 7257 (86%) | 556 (6.6%) | 506 (6%) | 87 (1%) | 35 (0.4%) |
| T2,MH,HF | 4763 | 69.8 [60.9, 78.2] | 658 (13.8%) | 774 (16.3%) | 921 (19.3%) | 1062 (22.3%) | 1348 (28.3%) | 2346 (49.3%) | 2417 (50.7%) | 4249 (89.2%) | 258 (5.4%) | 194 (4.1%) | 46 (1%) | 16 (0.3%) |

| | | | | | | | | | | | | | | |
|---|---|---|---|---|---|---|---|---|---|---|---|---|---|---|
| CK,MH,HF | 6247 | 80.5 [72.6, 86.2] | 1189 (19%) | 1315 (21.1%) | 1279 (20.5%) | 1220 (19.5%) | 1244 (19.9%) | 1913 (30.6%) | 4334 (69.4%) | 6033 (96.6%) | 109 (1.7%) | 80 (1.3%) | 13 (0.2%) | 12 (0.2%) |
| CV,T2,CK,MH | 18628 | 76 [69.3, 82.2] | 2959 (15.9%) | 3354 (18%) | 3577 (19.2%) | 4052 (21.8%) | 4686 (25.2%) | 8057 (43.3%) | 10571 (56.7%) | 16827 (90.3%) | 1137 (6.1%) | 465 (2.5%) | 144 (0.8%) | 55 (0.3%) |
| CV,T2,CK,HF | 14282 | 78.8 [72.6, 84.2] | 2295 (16.1%) | 2585 (18.1%) | 2877 (20.1%) | 3024 (21.2%) | 3501 (24.5%) | 9110 (63.8%) | 5172 (36.2%) | 12380 (86.7%) | 1236 (8.7%) | 499 (3.5%) | 109 (0.8%) | 58 (0.4%) |
| CV,T2,MH,HF | 6494 | 71.9 [63.9, 79.1] | 851 (13.1%) | 1045 (16.1%) | 1238 (19.1%) | 1491 (23%) | 1869 (28.8%) | 3952 (60.9%) | 2542 (39.1%) | 5817 (89.6%) | 462 (7.1%) | 143 (2.2%) | 43 (0.7%) | 29 (0.4%) |
| CV,CK,MH,HF | 9291 | 80.8 [74.2, 86.2] | 1627 (17.5%) | 1795 (19.3%) | 1873 (20.2%) | 1881 (20.2%) | 2115 (22.8%) | 4162 (44.8%) | 5129 (55.2%) | 8977 (96.6%) | 189 (2%) | 79 (0.9%) | 28 (0.3%) | 18 (0.2%) |
| T2,CK,MH,HF | 3244 | 77.2 [69.8, 83.5] | 489 (15.1%) | 589 (18.2%) | 630 (19.4%) | 712 (21.9%) | 824 (25.4%) | 1224 (37.7%) | 2020 (62.3%) | 2918 (90%) | 162 (5%) | 117 (3.6%) | 30 (0.9%) | 17 (0.5%) |
| All diseases | 6162 | 78 [71, 83.6] | 860 (14%) | 1075 (17.4%) | 1196 (19.4%) | 1381 (22.4%) | 1650 (26.8%) | 3087 (50.1%) | 3075 (49.9%) | 5548 (90%) | 387 (6.3%) | 156 (2.5%) | 51 (0.8%) | 20 (0.3%) |

* Age represents age at entry into the disease state.

Abbreviations: CV: cardiovascular disease; T2: type-2 diabetes; CK: chronic kidney disease; HF: heart failure; MH: mental health conditions.

Supplemental Table B. Summary of disease transitions.

**Summary of CVD diagnosis grouped by comorbidity**

| Previous Comorbidity | | Incidence Rate of CVD | Number of Events |
|---|---|---|---|
| 4 conditions | T2D,CKD,MH,HF | 0.059 | 521 |
| 3 conditions | CKD,MH,HF | 0.052 | 960 |
| | T2D,MH,HF | 0.061 | 741 |
| | T2D,CKD,HF | 0.058 | 1416 |
| | T2D,CKD,MH | 0.025 | 3370 |
| 2 conditions | MH,HF | 0.06 | 2313 |
| | CKD,HF | 0.056 | 2869 |
| | CKD,MH | 0.02 | 8343 |
| | T2D,HF | 0.063 | 1788 |
| | T2D,MH | 0.014 | 8677 |
| | T2D,CKD | 0.023 | 8525 |
| 1 condition | HF | 0.063 | 6047 |
| | MH | 0.004 | 18165 |
| | CKD | 0.02 | 23075 |
| | T2D | 0.012 | 15838 |
| 0 condition | NONE | 0.00046 | 207,102 |

Abbreviations: CVD: cardiovascular disease; T2D: type-2 diabetes; CKD: chronic kidney disease; HF: heart failure; MH: mental health conditions.

**Summary of T2D diagnosis grouped by comorbidity**

| Previous Comorbidity | | Incidence Rate of T2D | Number of Events |
|---|---|---|---|
| 4 conditions | CVD,CKD,MH,HF | 0.023 | 720 |
| 3 conditions | CKD,MH,HF | 0.02 | 375 |
| | CVD,MH,HF | 0.025 | 1101 |
| | CVD,CKD,HF | 0.022 | 1790 |
| | CVD,CKD,MH | 0.017 | 3111 |

| | | | |
|---|---|---|---|
| 2 conditions | MH,HF | 0.023 | 871 |
| | CKD,HF | 0.02 | 1012 |
| | CKD,MH | 0.013 | 5706 |
| | CVD,HF | 0.024 | 2362 |
| | CVD,MH | 0.019 | 9036 |
| | CVD,CKD | 0.016 | 7512 |
| 1 condition | HF | 0.021 | 2012 |
| | MH | 0.0055 | 25540 |
| | CKD | 0.013 | 15174 |
| | CVD | 0.017 | 16224 |
| 0 condition | NONE | 0.0006 | 269,791 |

Abbreviations: CVD: cardiovascular disease; T2D: type-2 diabetes; CKD: chronic kidney disease; HF: heart failure; MH: mental health conditions.

**Summary of CKD diagnosis grouped by comorbidity**

| Previous Comorbidity | | Incidence Rate of CKD | Number of Events |
|---|---|---|---|
| 4 conditions | CVD,T2D,MH,HF | 0.071 | 1449 |
| 3 conditions | T2D,MH,HF | 0.066 | 812 |
| | CVD,MH,HF | 0.054 | 2332 |
| | CVD,T2D,HF | 0.087 | 3703 |
| | CVD,T2D,MH | 0.03 | 5101 |
| 2 conditions | MH,HF | 0.048 | 1844 |
| | T2D,HF | 0.081 | 2306 |
| | T2D,MH | 0.014 | 8813 |
| | CVD,HF | 0.062 | 6117 |
| | CVD,MH | 0.021 | 10103 |
| | CVD,T2D | 0.034 | 11028 |
| 1 condition | HF | 0.056 | 5408 |
| | MH | 0.0029 | 13347 |

| | | | |
|---|---|---|---|
| | T2D | 0.014 | 19008 |
| | CVD | 0.023 | 21873 |
| 0 condition | NONE | 0.00047 | 211,219 |

Abbreviations: CVD: cardiovascular disease; T2D: type-2 diabetes; CKD: chronic kidney disease; HF: heart failure; MH: mental health conditions.

**Summary of MH diagnosis grouped by comorbidity**

| Previous Comorbidity | | Incidence Rate of MH | Number of Events |
|---|---|---|---|
| 4 conditions | CVD,T2D,CKD,HF | 0.015 | 702 |
| 3 conditions | T2D,CKD,HF | 0.013 | 316 |
| | CVD,CKD,HF | 0.014 | 1197 |
| | CVD,T2D,HF | 0.016 | 674 |
| | CVD,T2D,CKD | 0.013 | 2488 |
| 2 conditions | CKD,HF | 0.012 | 635 |
| | T2D,HF | 0.016 | 466 |
| | T2D.CKD | 0.011 | 4093 |
| | CVD,HF | 0.016 | 1526 |
| | CVD,CKD | 0.013 | 5780 |
| | CVD,T2D | 0.015 | 4943 |
| 1 condition | HF | 0.015 | 1425 |
| | CKD | 0.011 | 12476 |
| | T2D | 0.014 | 18600 |
| | CVD | 0.016 | 15087 |
| 0 condition | NONE | 0.002 | 947,978 |

Abbreviations: CVD: cardiovascular disease; T2D: type-2 diabetes; CKD: chronic kidney disease; HF: heart failure; MH: mental health conditions.

**Summary of HF diagnosis grouped by comorbidity**

| Previous Comorbidity | | Incidence Rate of HF | Number of Events |
|---|---|---|---|
| 4 conditions | CVD,T2D,CKD,MH | 0.024 | 1924 |
| 3 conditions | T2D,CKD,MH | 0.01 | 1321 |
| | CVD,CKD,MH | 0.017 | 3054 |
| | CVD,T2D,MH | 0.012 | 2089 |
| | CVD,T2D,CKD | 0.024 | 4664 |
| 2 conditions | CKD,MH | 0.0062 | 2644 |
| | T2D,MH | 0.0031 | 1928 |
| | T2D,CKD | 0.0098 | 3631 |
| | CVD,MH | 0.0088 | 4242 |
| | CVD,CKD | 0.018 | 8085 |
| | CVD,T2D | 0.013 | 4304 |
| 1 condition | MH | 0.00071 | 3310 |
| | CKD | 0.0066 | 7707 |
| | T2D | 0.0027 | 3660 |
| | CVD | 0.0095 | 8868 |
| 0 condition | NONE | 0.000079 | 35434 |

Abbreviations: CVD: cardiovascular disease; T2D: type-2 diabetes; CKD: chronic kidney disease; HF: heart failure; MH: mental health conditions.

Supplemental Figure B. Plots of the estimated cumulative hazard of the Cox-Snell residuals.

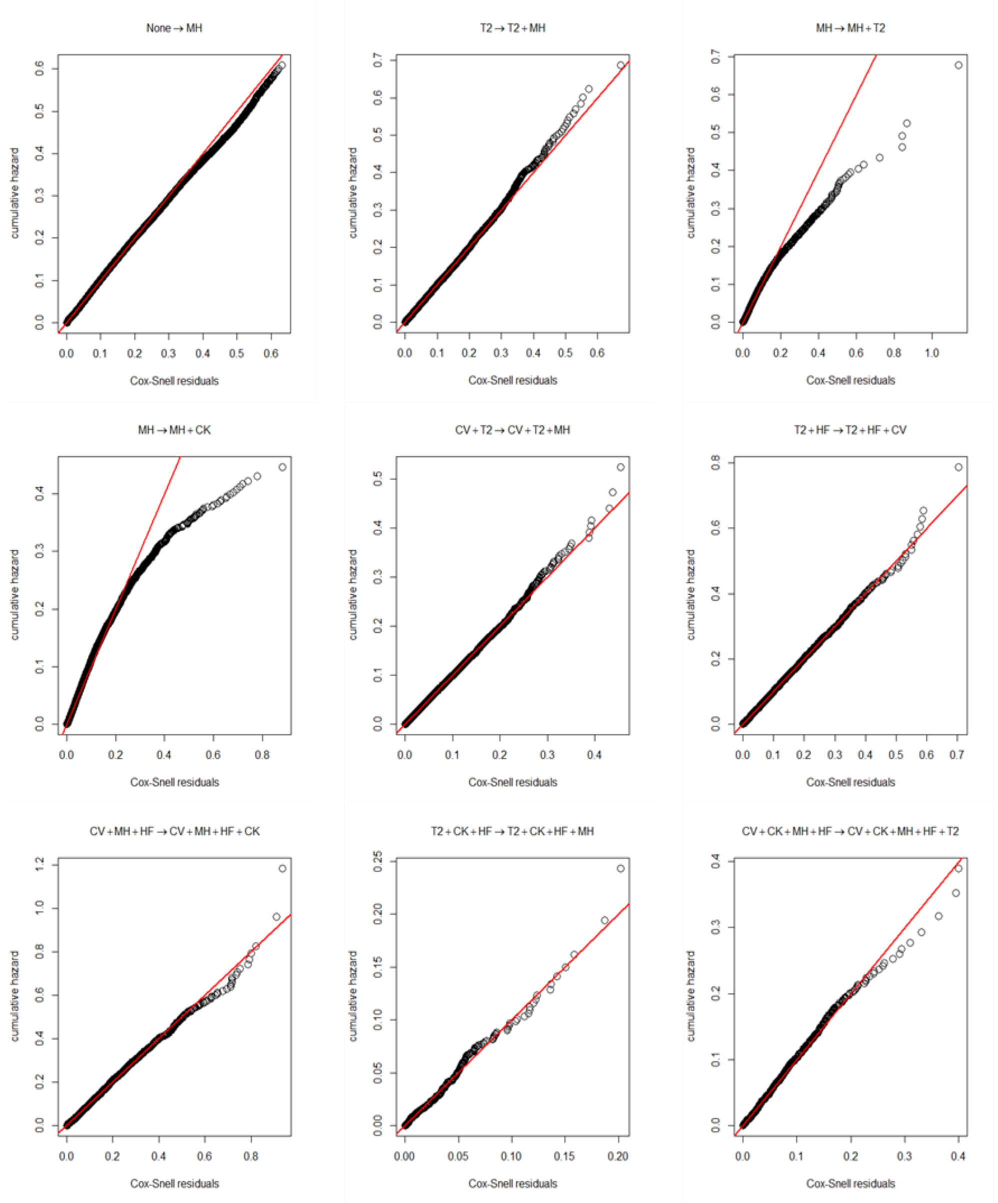


Note: The red line is a reference line with an intercept of zero and a slope of one. Abbreviations: CV: cardiovascular disease; T2: type-2 diabetes; CK: chronic kidney disease; HF: heart failure; MH: mental health conditions.

#### Supplemental Figure C. Estimated association between each sociodemographic characteristic and the rate of CVD or MH diagnosis based on their respective narrower definitions

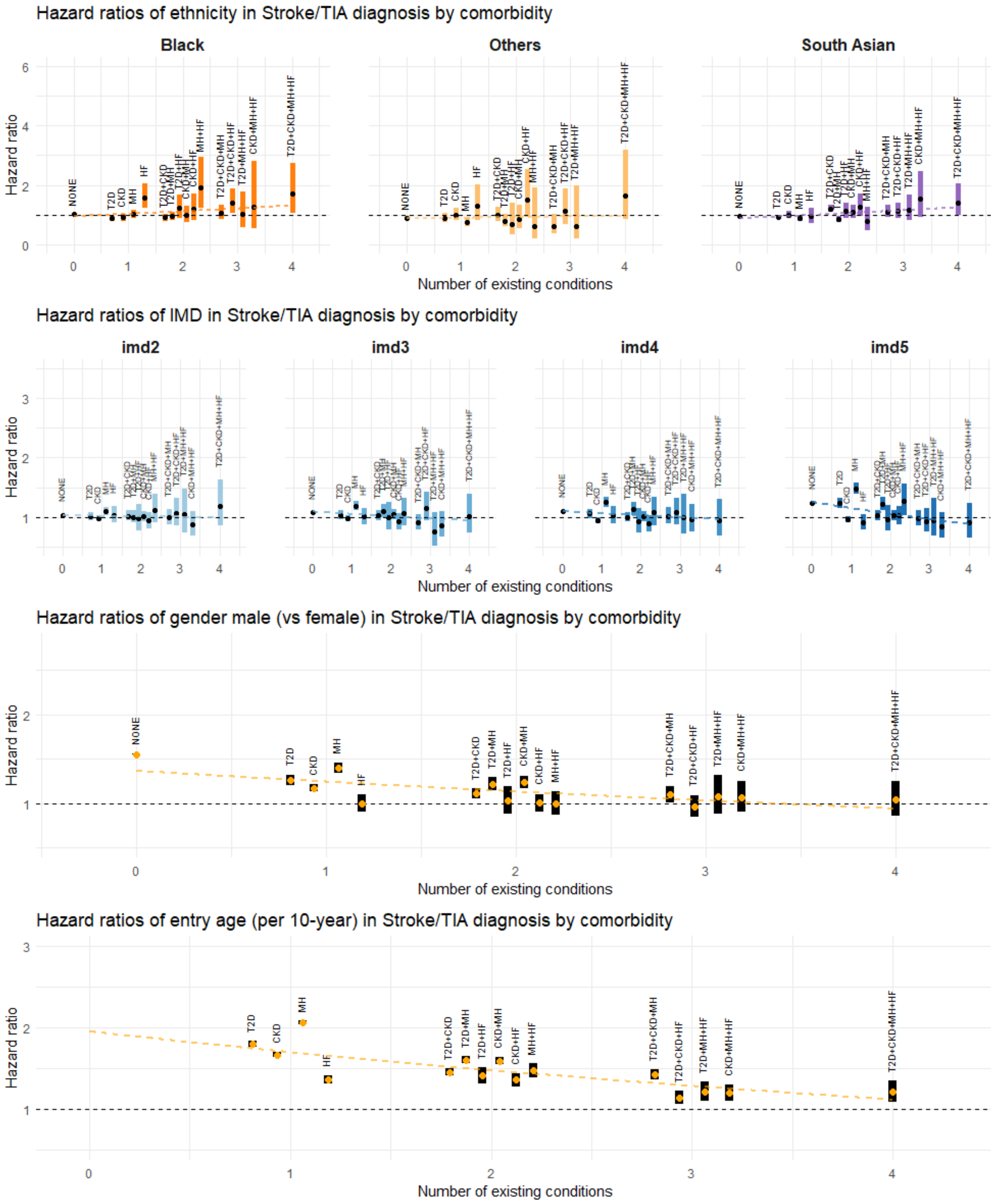


Figure C.1. Estimated association between each sociodemographic characteristic and the rate of CVD diagnosis (defined based on stroke/TIA only). Graphical settings are the same as in Figures 2-5 of the main paper. Abbreviations: CVD: cardiovascular disease; T2D: type-2 diabetes; CKD: chronic kidney disease; HF: heart failure; MH: mental health conditions; TIA: transient ischaemic attack.

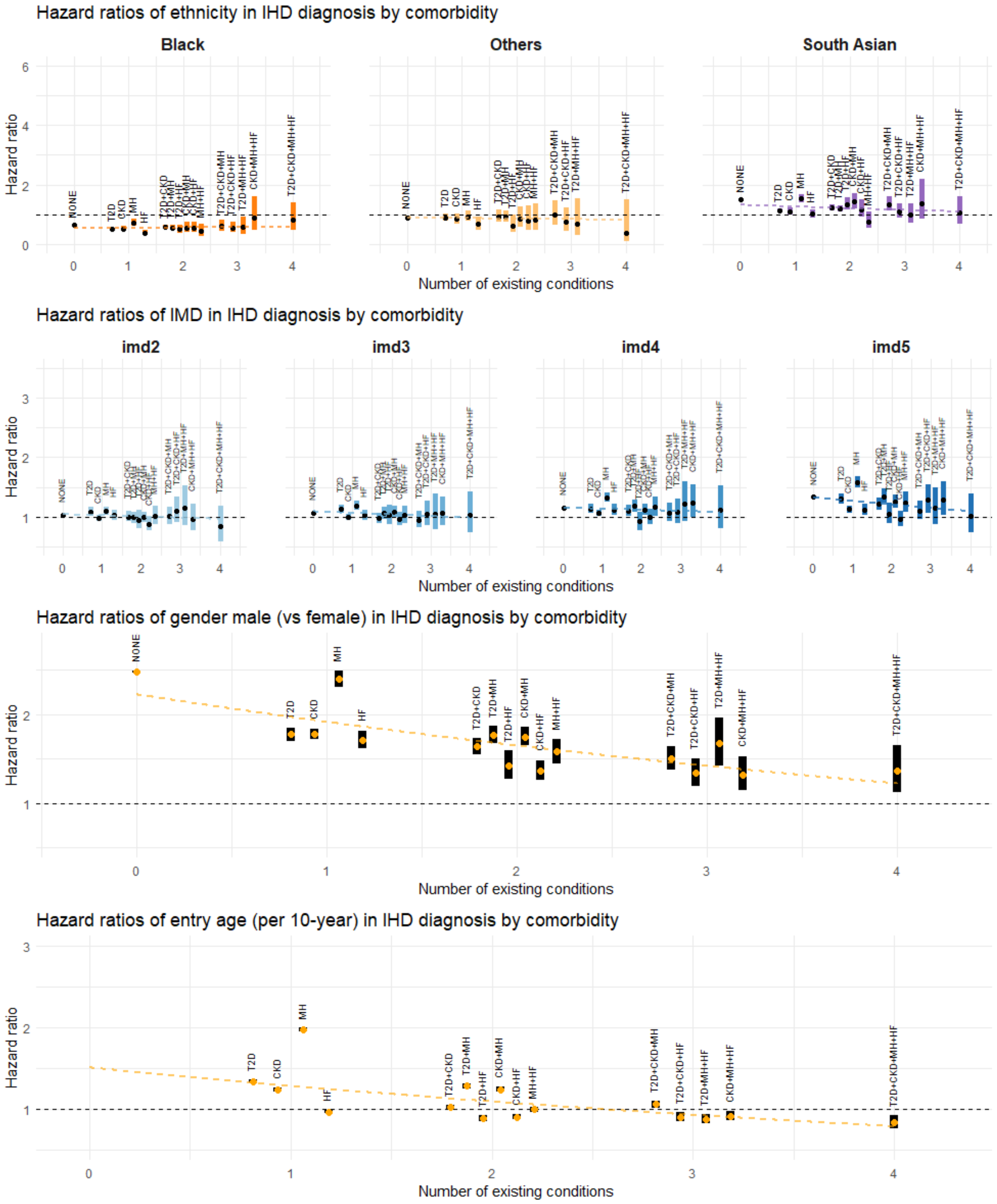


Figure C.2. Estimated association between each sociodemographic characteristic and the rate of CVD diagnosis (defined based on IHD only). Graphical settings are the same as in Figures 2-5 of the main paper. Abbreviations: CVD: cardiovascular disease; T2D: type-2 diabetes; CKD: chronic kidney disease; HF: heart failure; MH: mental health conditions; IHD: ischaemic heart disease.

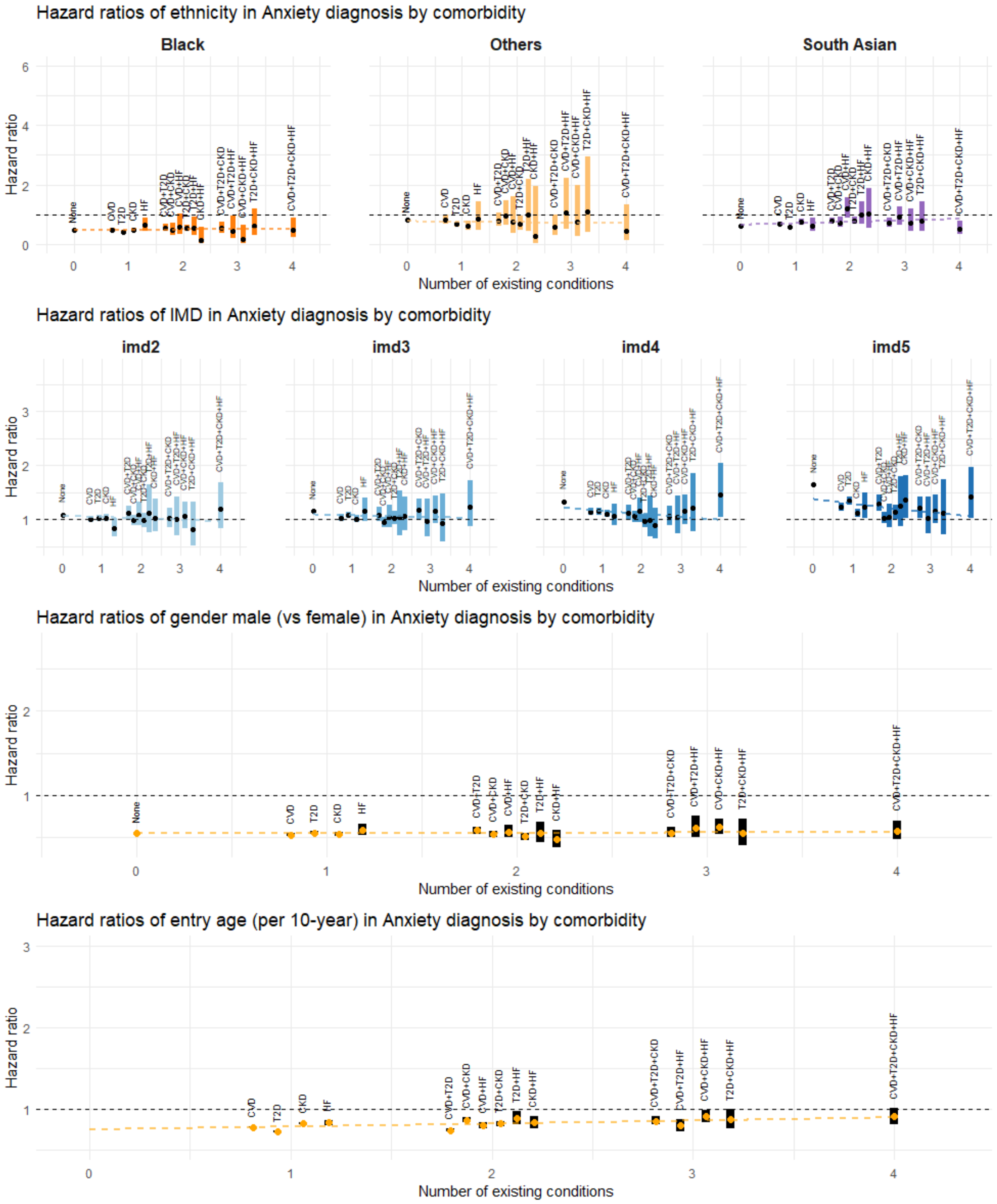


Figure C.3. Estimated association between each sociodemographic characteristic and the rate of MH diagnosis (defined based on anxiety only). Graphical settings are the same as in Figures 2-5 of the main paper. Abbreviations: CVD: cardiovascular disease; T2D: type-2 diabetes; CKD: chronic kidney disease; HF: heart failure; MH: mental health conditions.

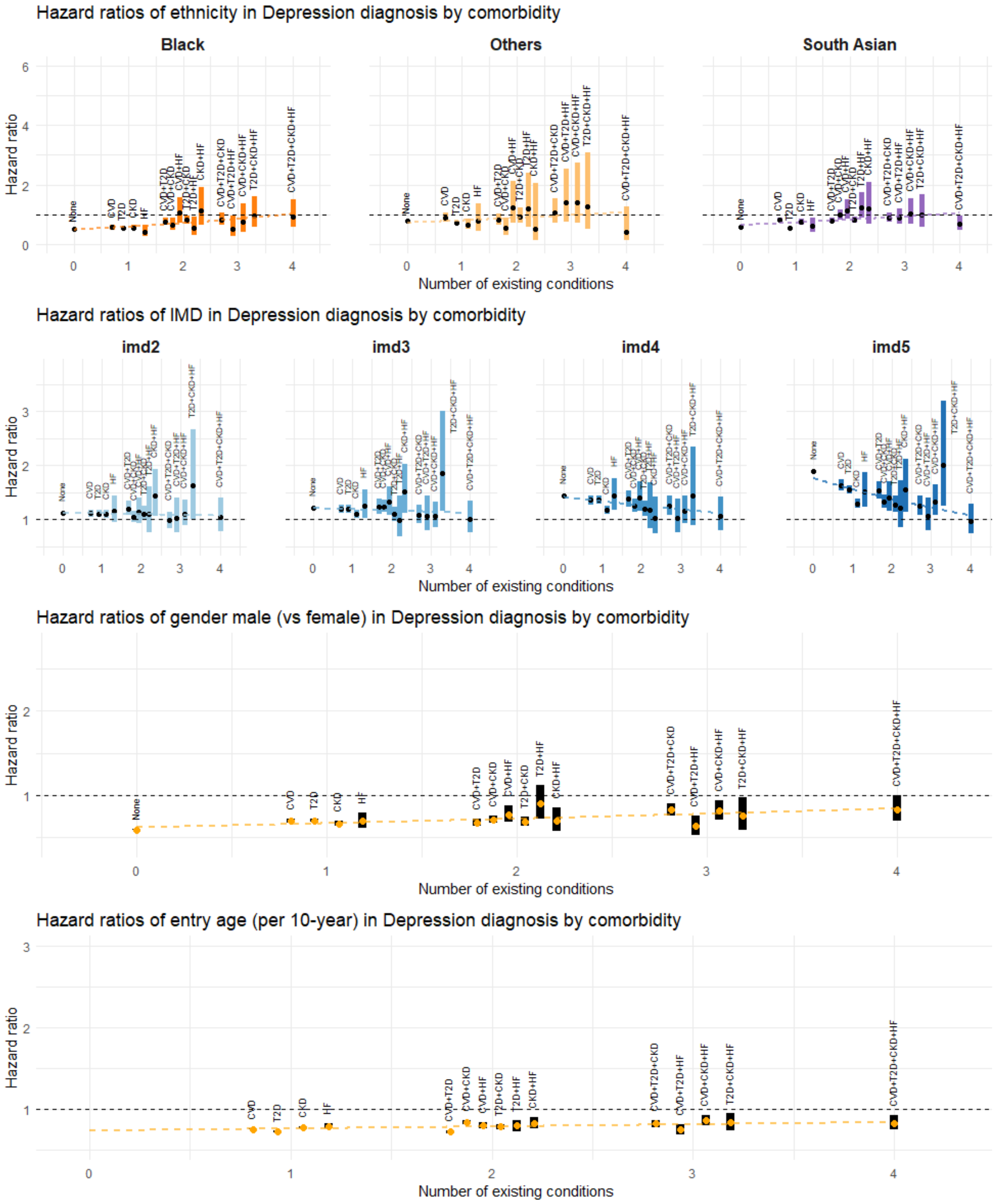


Figure C.4. Estimated association between each sociodemographic characteristic and the rate of MH diagnosis (defined based on depression only). Graphical settings are the same as in Figures 2-5 of the main paper. Abbreviations: CVD: cardiovascular disease; T2D: type-2 diabetes; CKD: chronic kidney disease; HF: heart failure; MH: mental health conditions.

Supplemental Figure D. Results of the multistate analysis obtained with COPD replacing HF, focusing on the associations between each sociodemographic characteristic and the rate of CVD, T2D, CKD, or MH diagnosis.

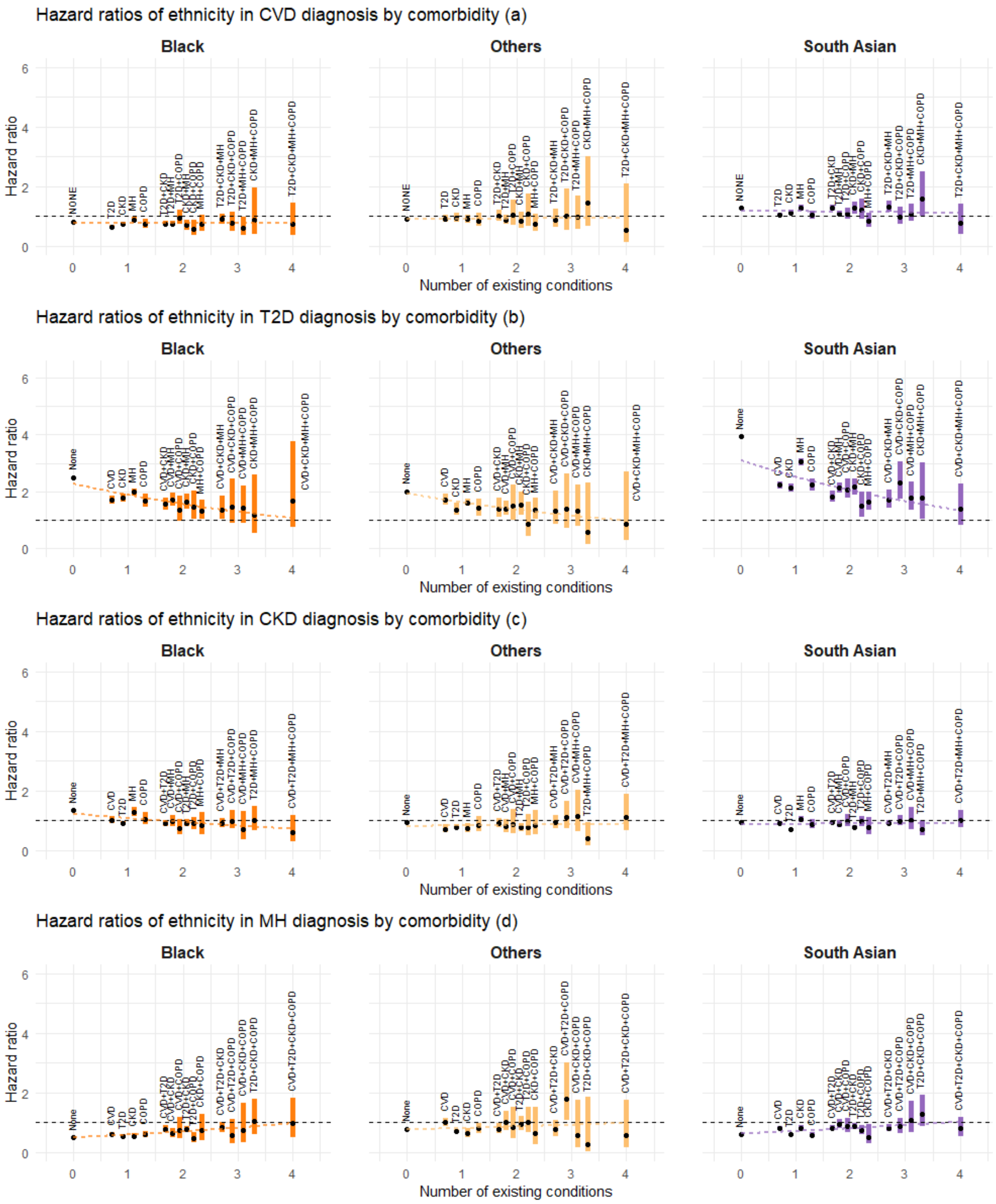


Figure D.1. Estimated association between ethnicity (groups) and the rate of disease transition by number of existing conditions and comorbidity status. Graphical settings are the same as in Figure 2 of the main paper. Abbreviations: CVD: cardiovascular disease; T2D: type-2 diabetes; CKD: chronic kidney disease; HF: heart failure; MH: mental health conditions; COPD: chronic obstructive pulmonary disease.

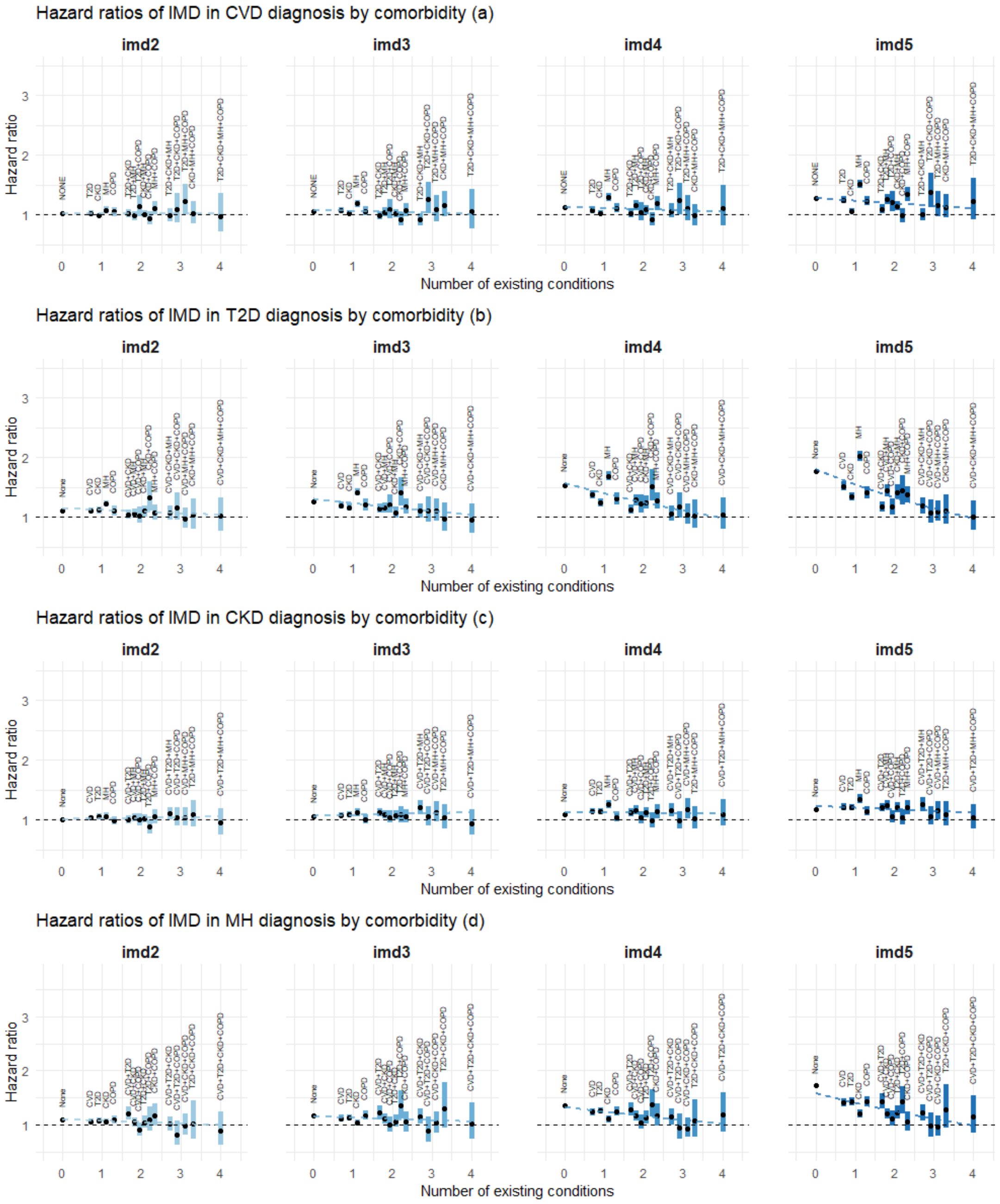


Figure D.2. Estimated association between (IMD quintiles 2-5) and the rate of disease transition by number of existing conditions and comorbidity status. Graphical settings are the same as in Figure 3 of the main paper. Abbreviations: CVD: cardiovascular disease; T2D: type-2 diabetes; CKD: chronic kidney disease; HF: heart failure; MH: mental health conditions; COPD: chronic obstructive pulmonary disease.

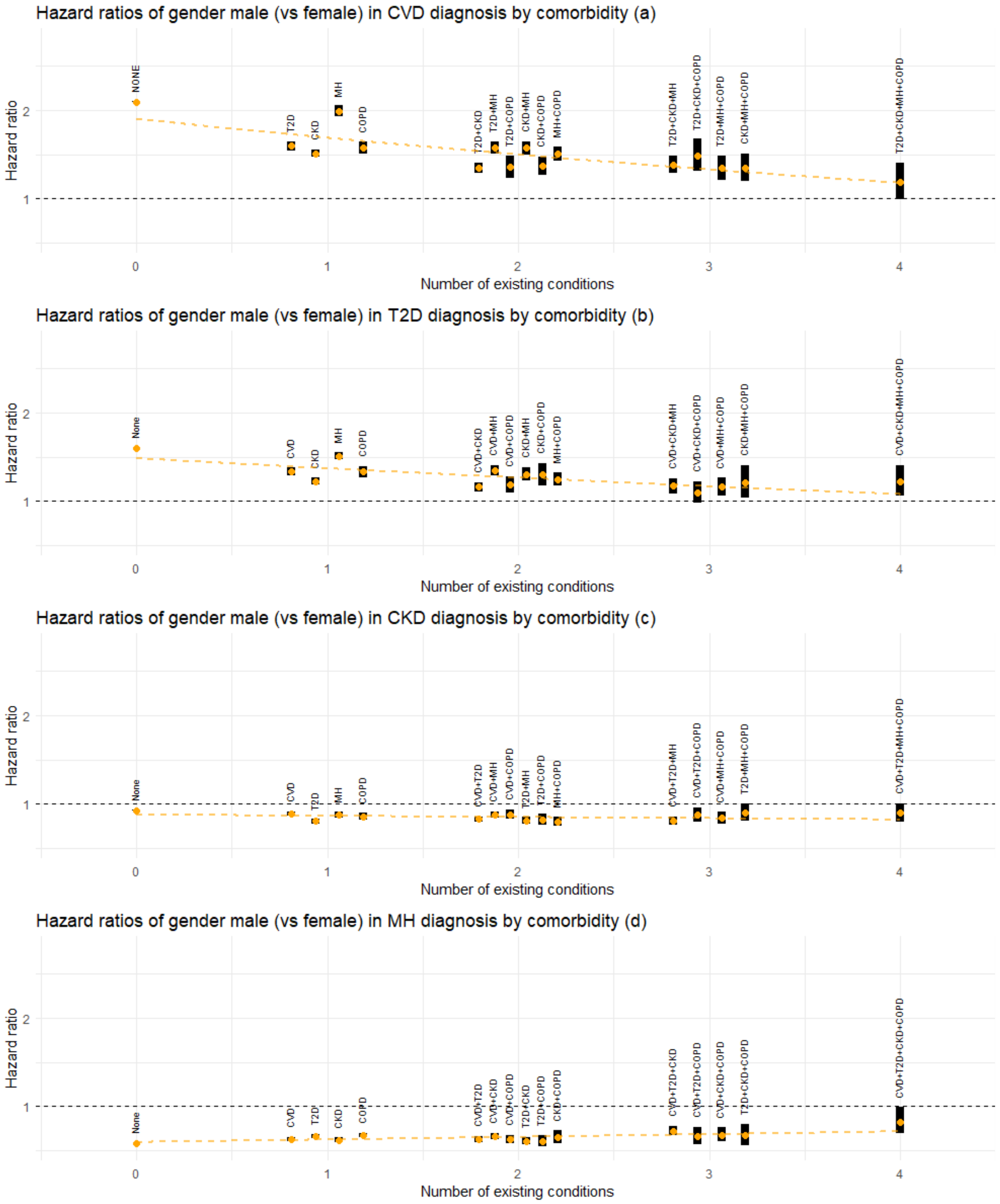


Figure D.3. Estimated association between gender (male) and the rate of disease transition by number of existing conditions and comorbidity status. Graphical settings are the same as in Figure 4 of the main paper. Abbreviations: CVD: cardiovascular disease; T2D: type-2 diabetes; CKD: chronic kidney disease; HF: heart failure; MH: mental health conditions; COPD: chronic obstructive pulmonary disease.

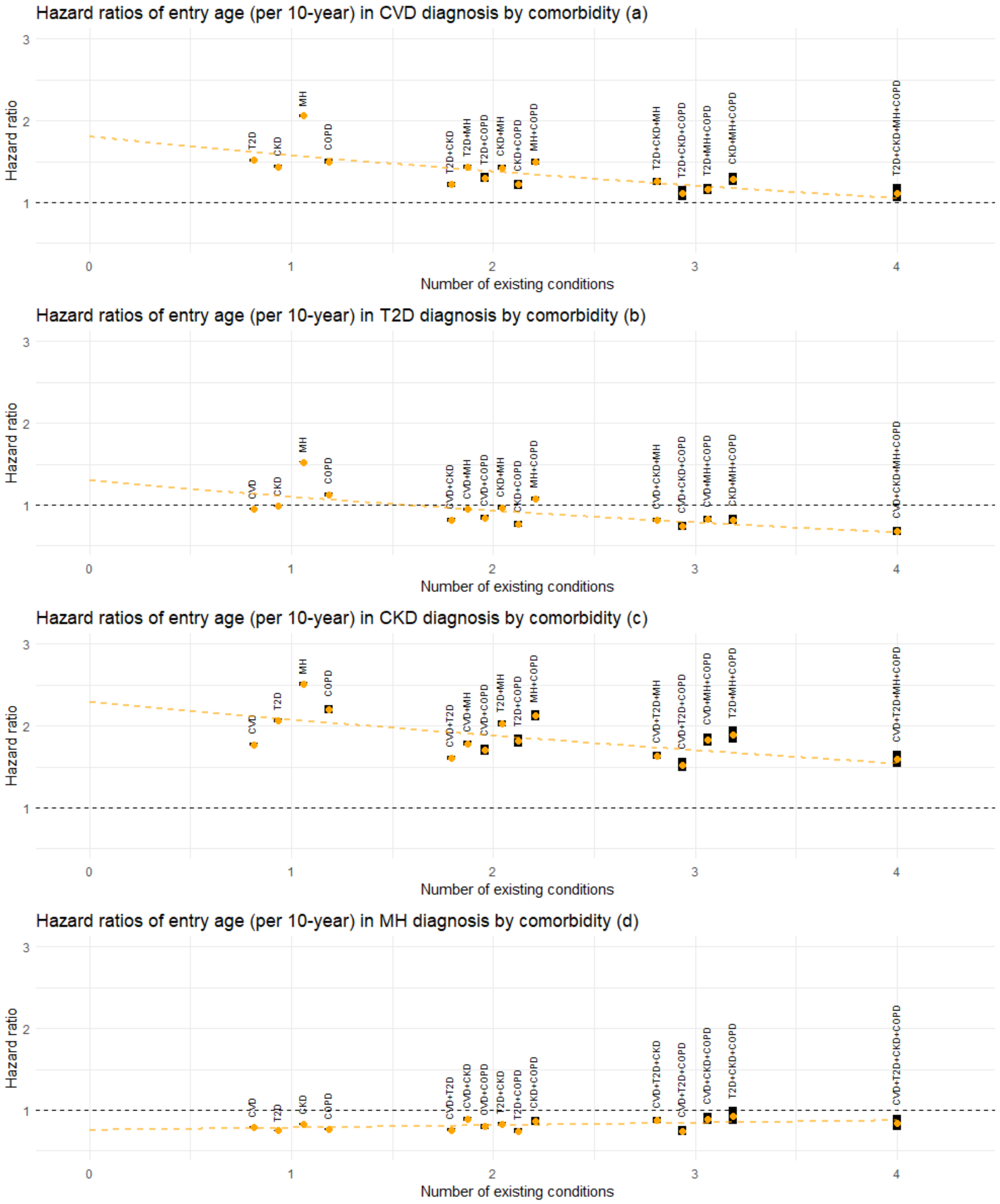


Figure D.4. Estimated association between age at entry to the current state (per 10 years) and the rate of next disease transition by number of existing conditions and comorbidity status. Graphical settings are the same as in Figure 5 of the main paper. Abbreviations: CVD: cardiovascular disease; T2D: type-2 diabetes; CKD: chronic kidney disease; HF: heart failure; MH: mental health conditions; COPD: chronic obstructive pulmonary disease.

Supplemental Figure E. Results of the multistate analysis with COPD included as the sixth condition in addition to CVD, T2D, CKD, HF and MH, focusing on the same transition types as in Figures 2-5.

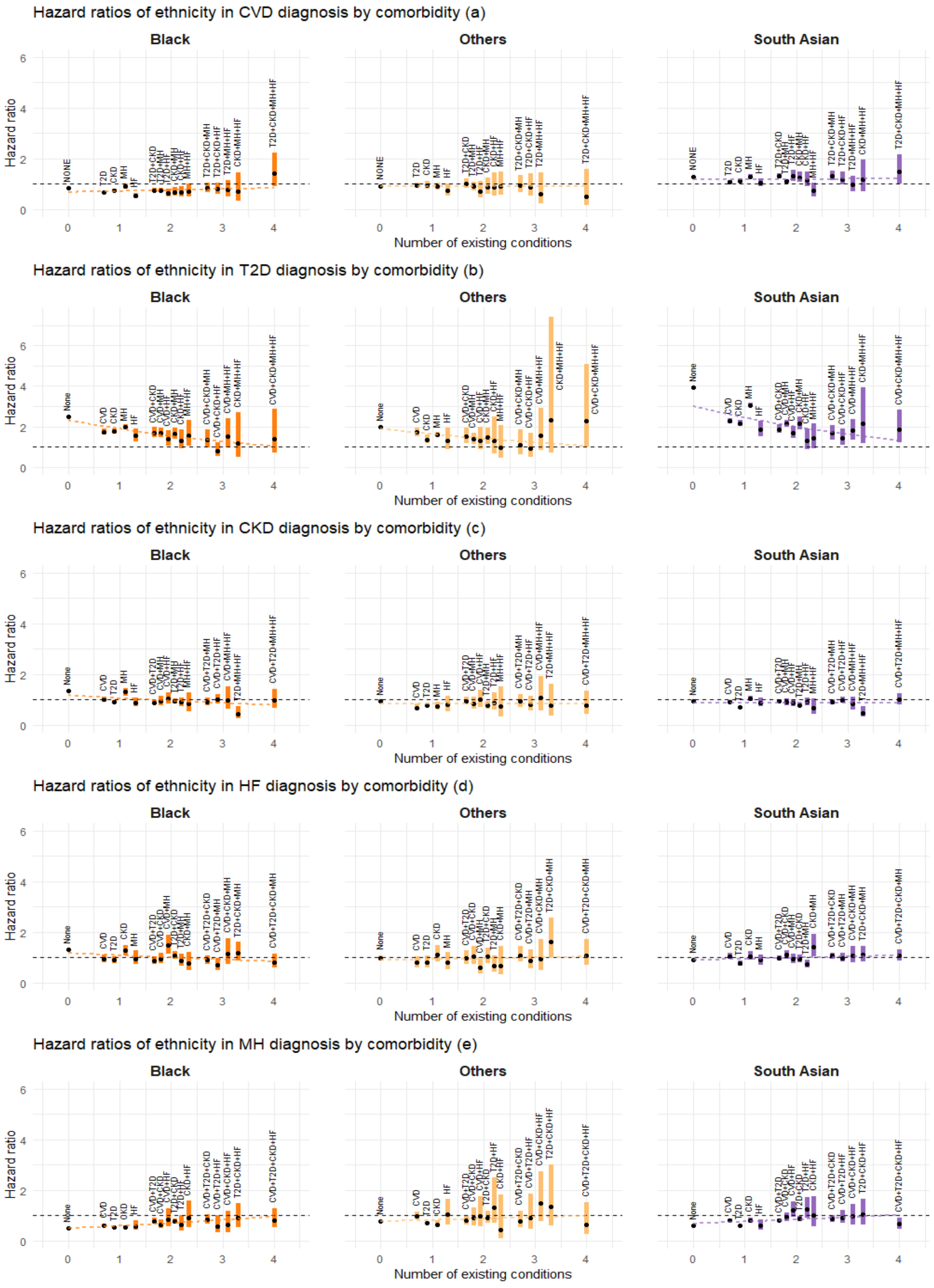


Figure E.1. Estimated association between ethnicity (groups) and the rate of disease transition by number of existing conditions and comorbidity status. Graphical settings are the same as in Figure 2 of the main paper. Abbreviations: CVD: cardiovascular disease; T2D: type-2 diabetes; CKD: chronic kidney disease; HF: heart failure; MH: mental health conditions; COPD: chronic obstructive pulmonary disease.

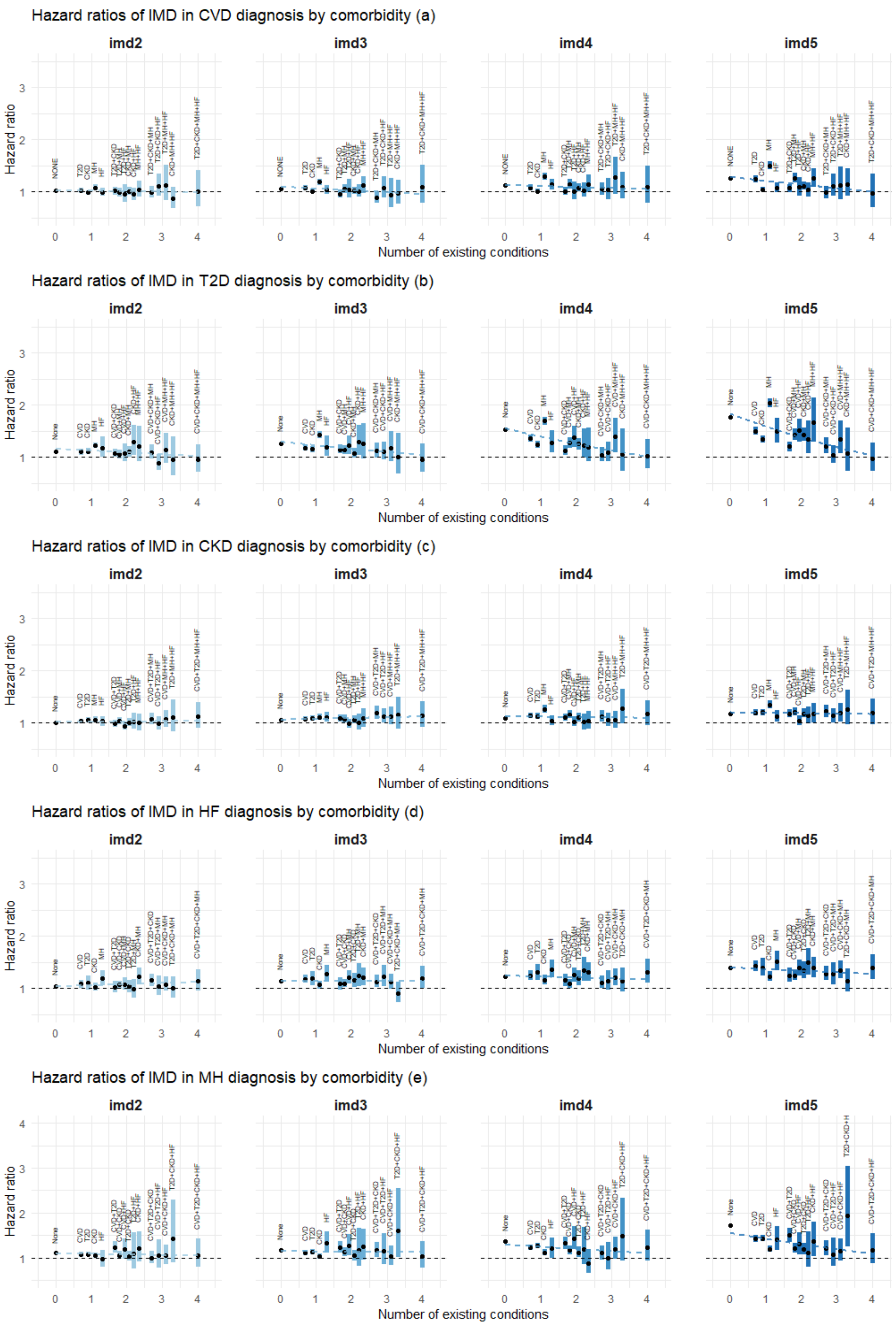


Figure E.2. Estimated association between (IMD quintiles 2-5) and the rate of disease transition by number of existing conditions and comorbidity status. Graphical settings are the same as in Figure 3 of the main paper. Abbreviations: CVD: cardiovascular disease; T2D: type-2 diabetes; CKD: chronic kidney disease; HF: heart failure; MH: mental health conditions; COPD: chronic obstructive pulmonary disease.

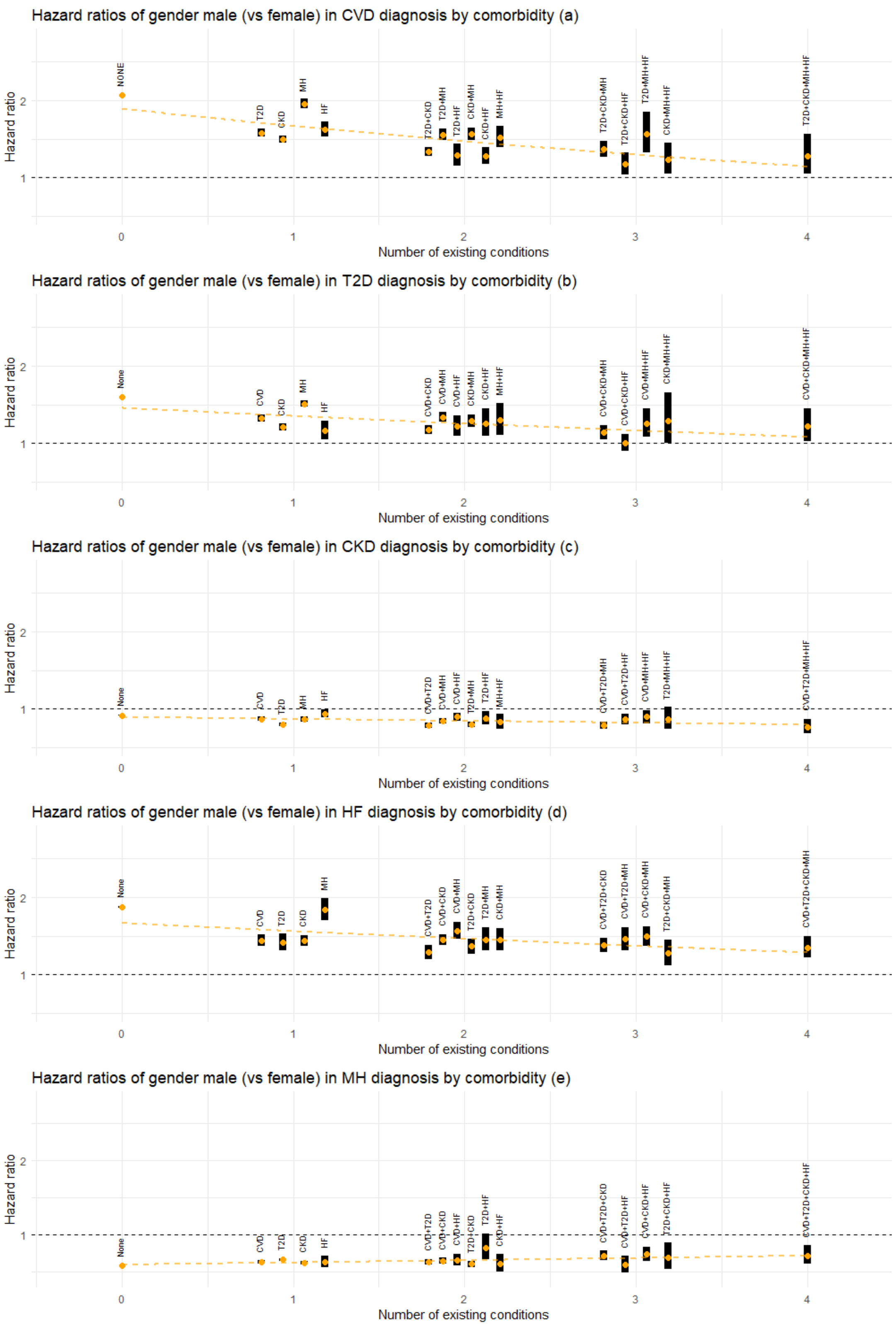


Figure E.3. Estimated association between gender (male) and the rate of disease transition by number of existing conditions and comorbidity status. Graphical settings are the same as in Figure 4 of the main paper. Abbreviations: CVD: cardiovascular disease; T2D: type-2 diabetes; CKD: chronic kidney disease; HF: heart failure; MH: mental health conditions; COPD: chronic obstructive pulmonary disease.

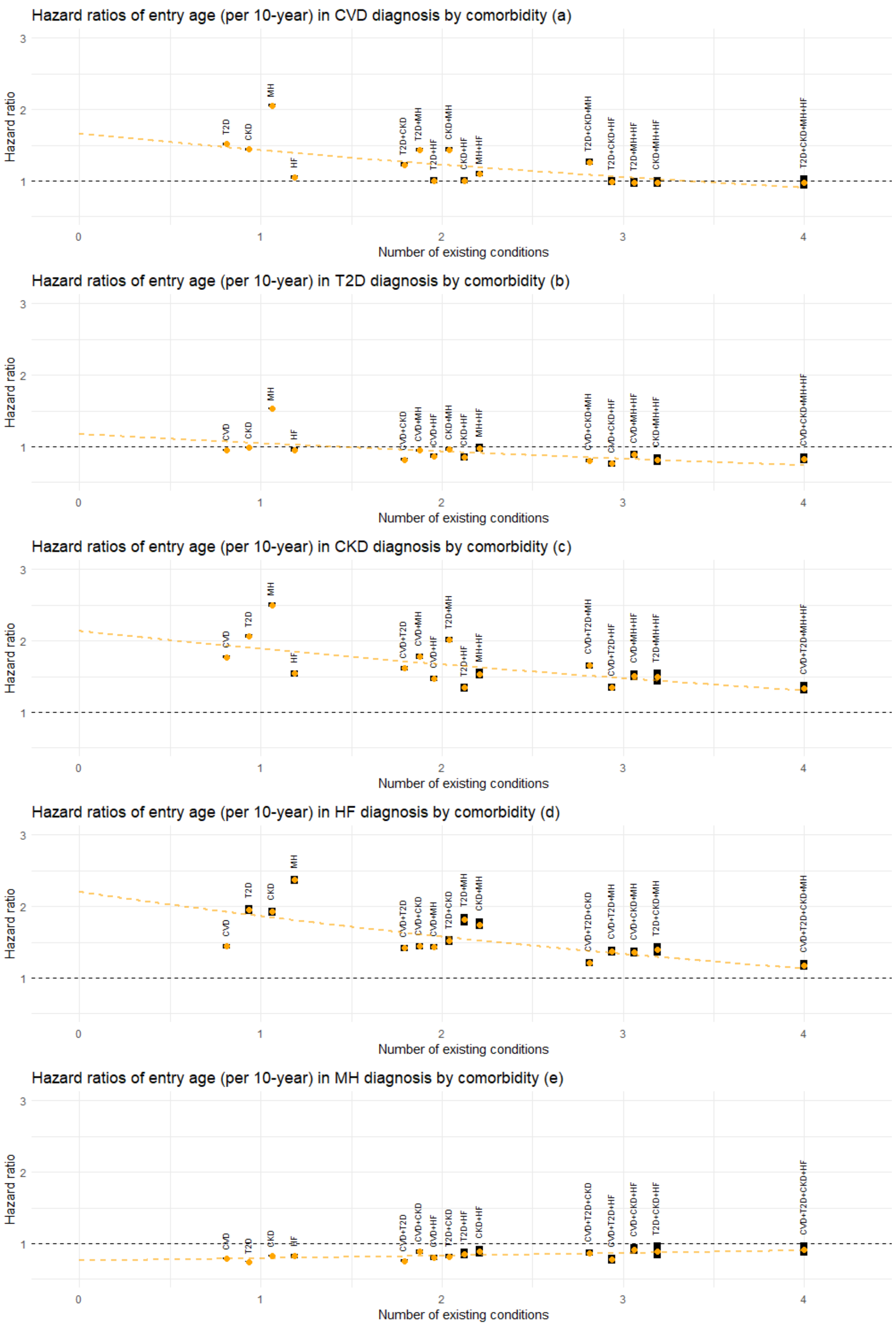


Figure E.4. Estimated association between age at entry to the current state (per 10 years) and the rate of next disease transition by number of existing conditions and comorbidity status. Graphical settings are the same as in Figure 5 of the main paper. Abbreviations: CVD: cardiovascular disease; T2D: type-2 diabetes; CKD: chronic kidney disease; HF: heart failure; MH: mental health conditions; COPD: chronic obstructive pulmonary disease.

Supplemental Figure F. Estimated association between deprivation (IMD deciles) and the rate of disease transition by number of existing conditions and comorbidity status. Panels from left to right show HRs for 2nd (left) – 10th (right) IMD deciles. The 1st IMD decile (the least deprived group) is treated as the reference category. For each transition and each category, the estimated HR is shown by an orange dot, with the corresponding 95% confidence interval represented by a black band.

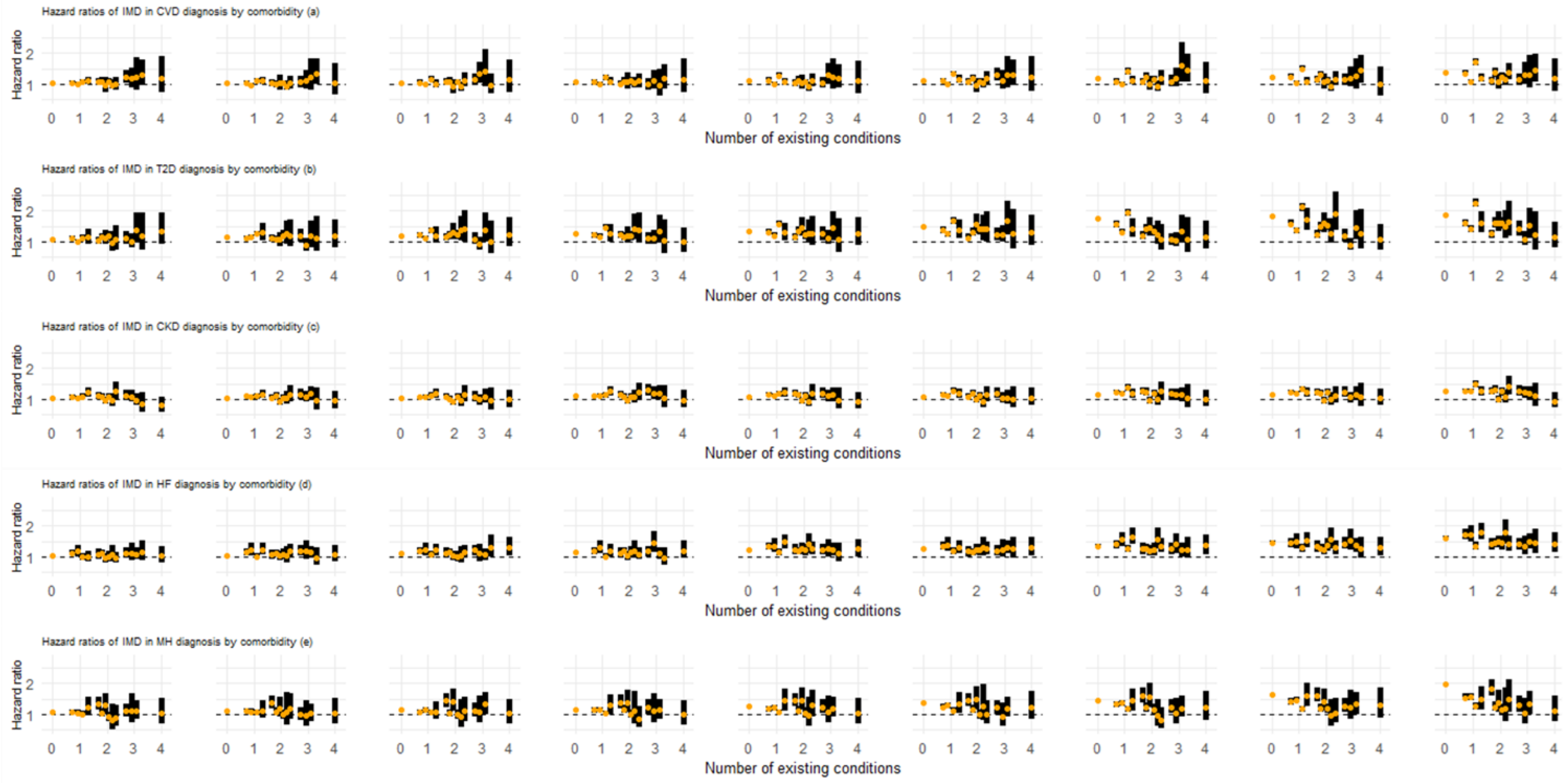

Supplemental Figure G. Results of the multistate analysis obtained based on the expanded cohort including patients with missing ethnicity information.

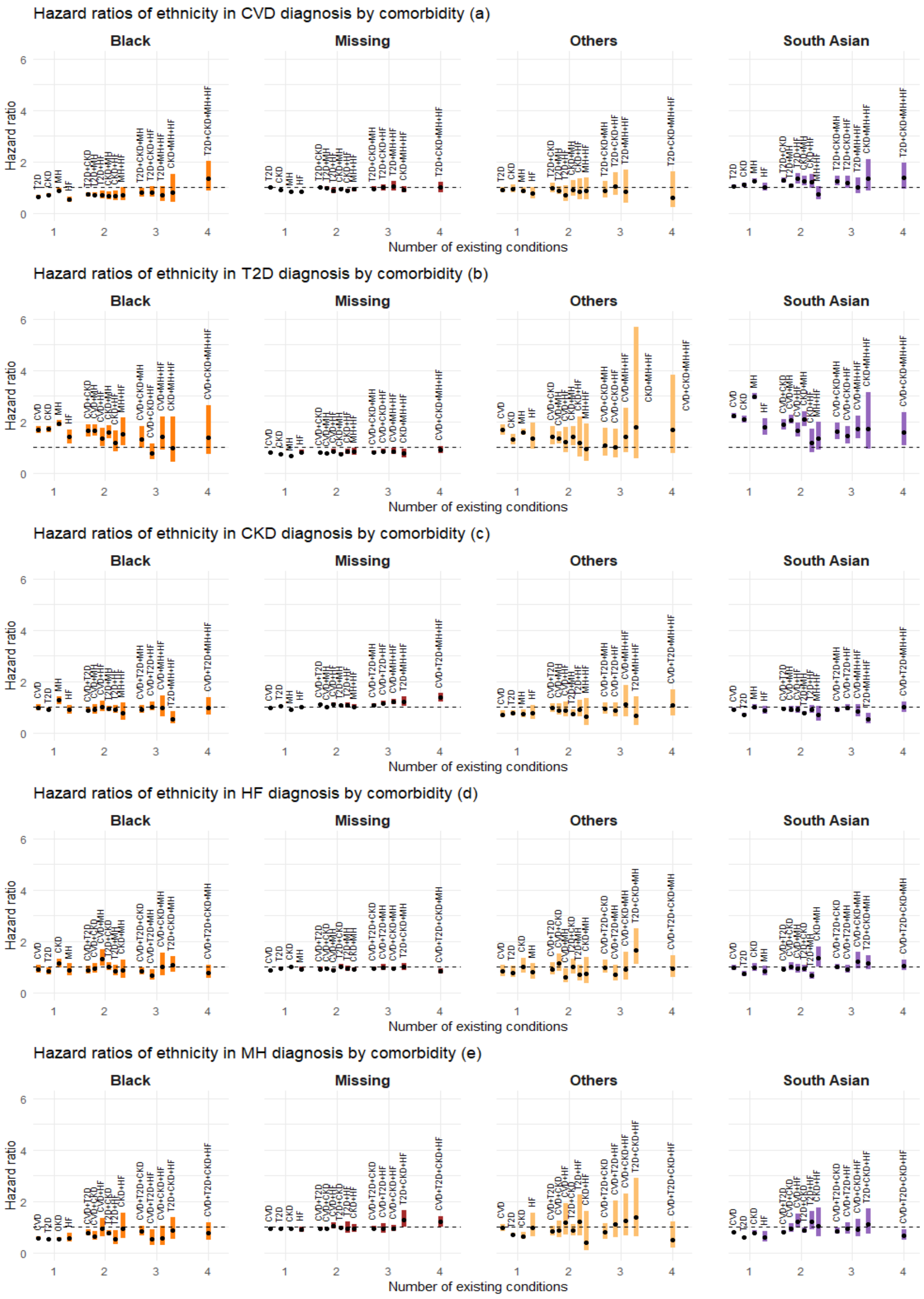


Figure G.1. Estimated association between ethnicity (groups) and the rate of disease transition by number of existing conditions and comorbidity status. Patients with missing ethnicity were assigned to the group “Missing”. Graphical settings are the same as in Figure 2 of the main paper. Abbreviations: CVD: cardiovascular disease; T2D: type-2 diabetes; CKD: chronic kidney disease; HF: heart failure; MH: mental health conditions.

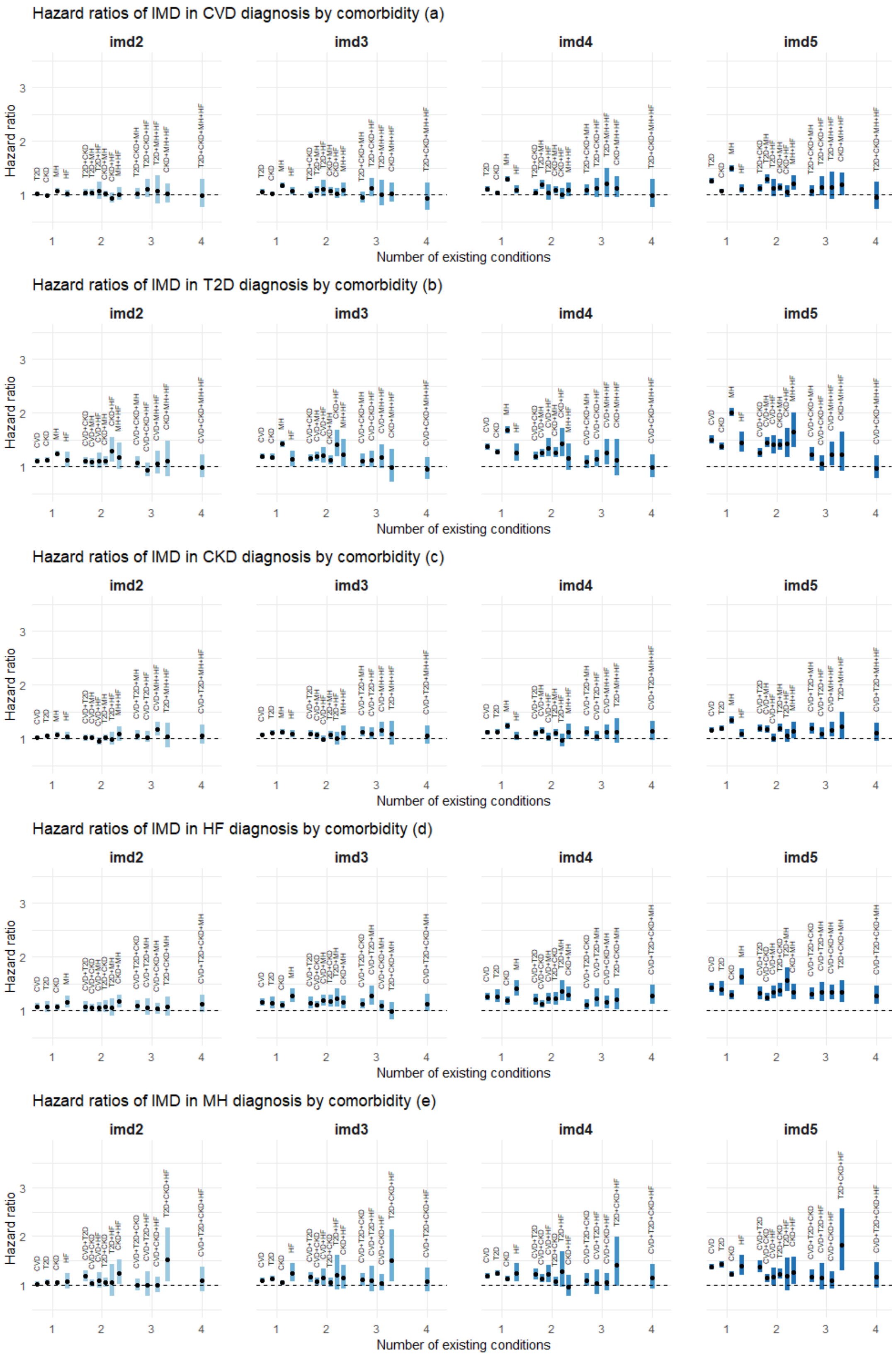


Figure G.2. Estimated association between (IMD quintiles 2-5) and the rate of disease transition by number of existing conditions and comorbidity status. Graphical settings are the same as in Figure 3 of the main paper. Abbreviations: CVD: cardiovascular disease; T2D: type-2 diabetes; CKD: chronic kidney disease; HF: heart failure; MH: mental health conditions.

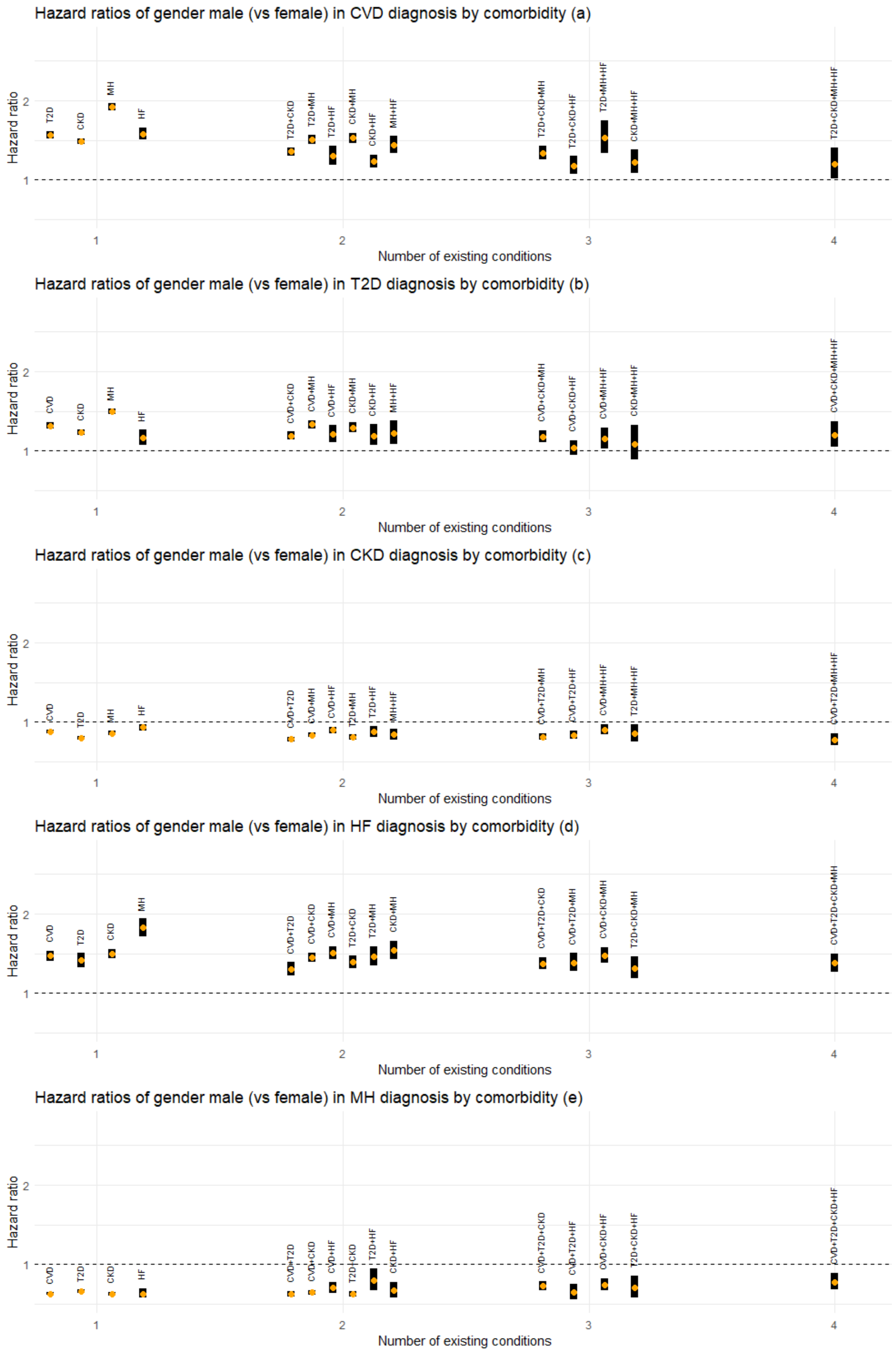


Figure G.3. Estimated association between gender (male) and the rate of disease transition by number of existing conditions and comorbidity status. Graphical settings are the same as in Figure 4 of the main paper. Abbreviations: CVD: cardiovascular disease; T2D: type-2 diabetes; CKD: chronic kidney disease; HF: heart failure; MH: mental health conditions.

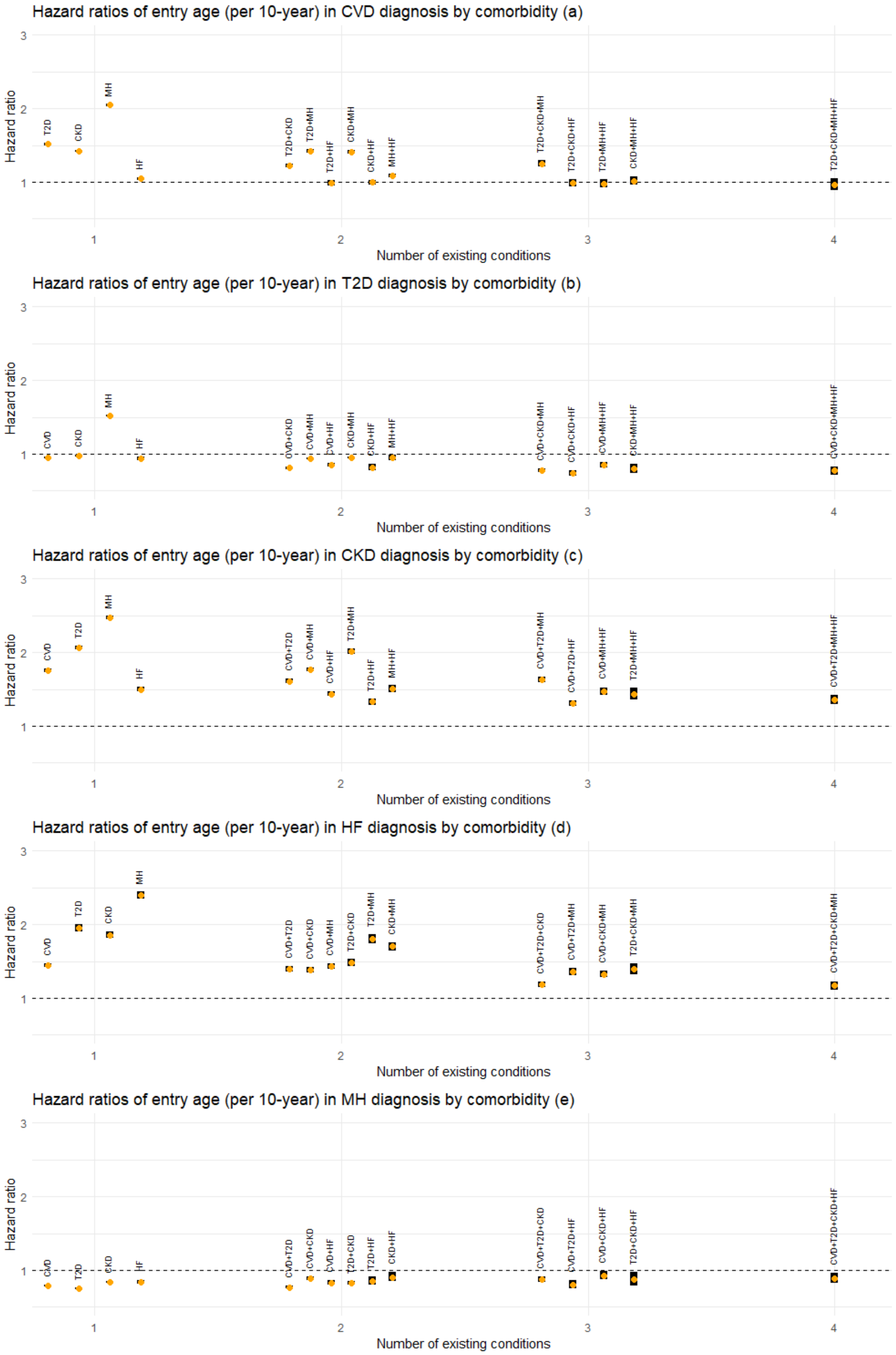


Figure G.4. Estimated association between age at entry to the current state (per 10 years) and the rate of next disease transition by number of existing conditions and comorbidity status. Graphical settings are the same as in Figure 5 of the main paper. Abbreviations: CVD: cardiovascular disease; T2D: type-2 diabetes; CKD: chronic kidney disease; HF: heart failure; MH: mental health conditions.

#### Supplemental Figure H. Results of the multistate analysis with transitions specified via semi-parametric Cox models.

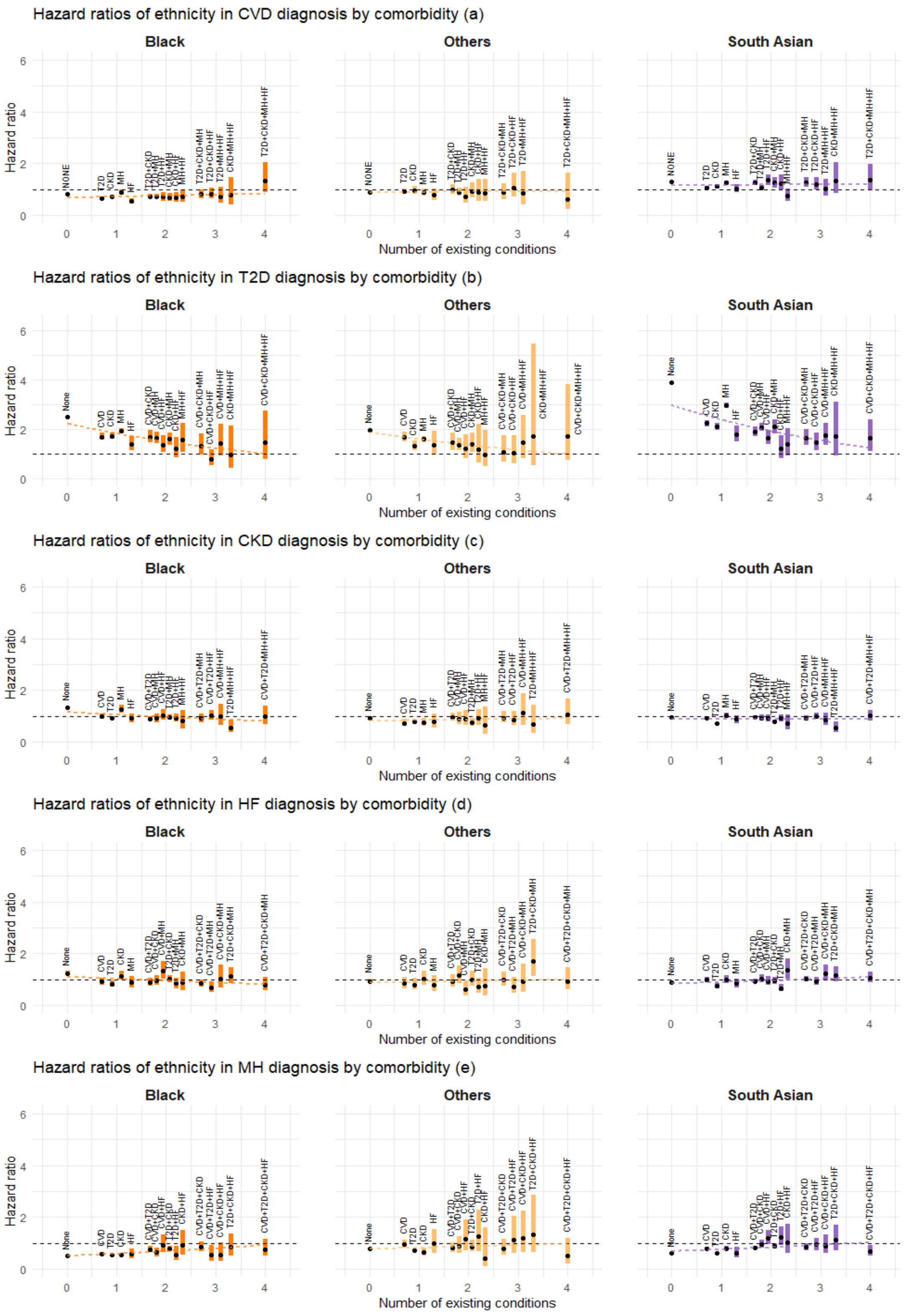


Figure H.1. Estimated association between ethnicity (groups) and the rate of disease transition by number of existing conditions and comorbidity status. Graphical settings are the same as in Figure 2 of the main paper. Abbreviations: CVD: cardiovascular disease; T2D: type-2 diabetes; CKD: chronic kidney disease; HF: heart failure; MH: mental health conditions.

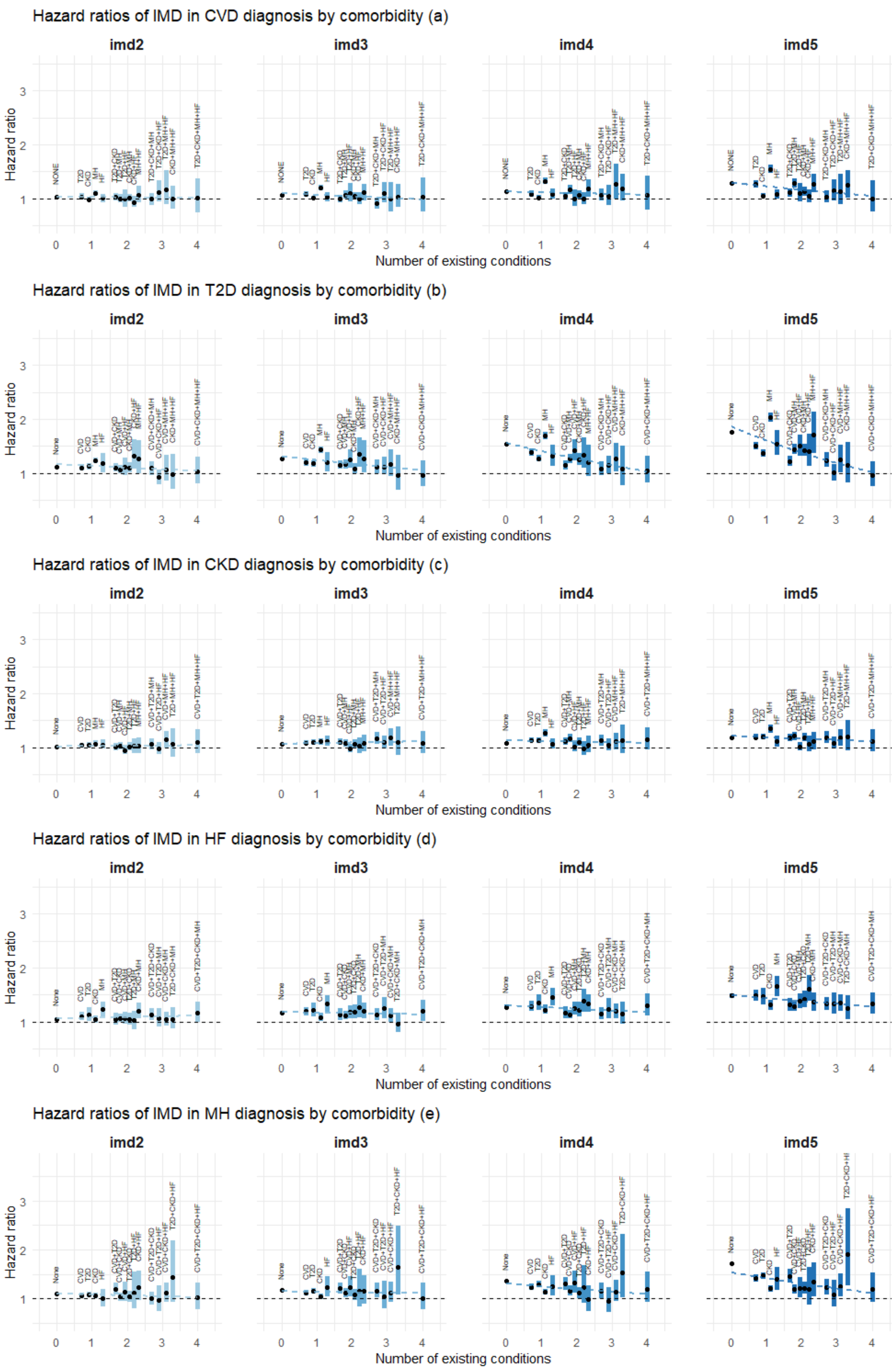


Figure H.2. Estimated association between (IMD quintiles 2-5) and the rate of disease transition by number of existing conditions and comorbidity status. Graphical settings are the same as in Figure 3 of the main paper. Abbreviations: CVD: cardiovascular disease; T2D: type-2 diabetes; CKD: chronic kidney disease; HF: heart failure; MH: mental health conditions.

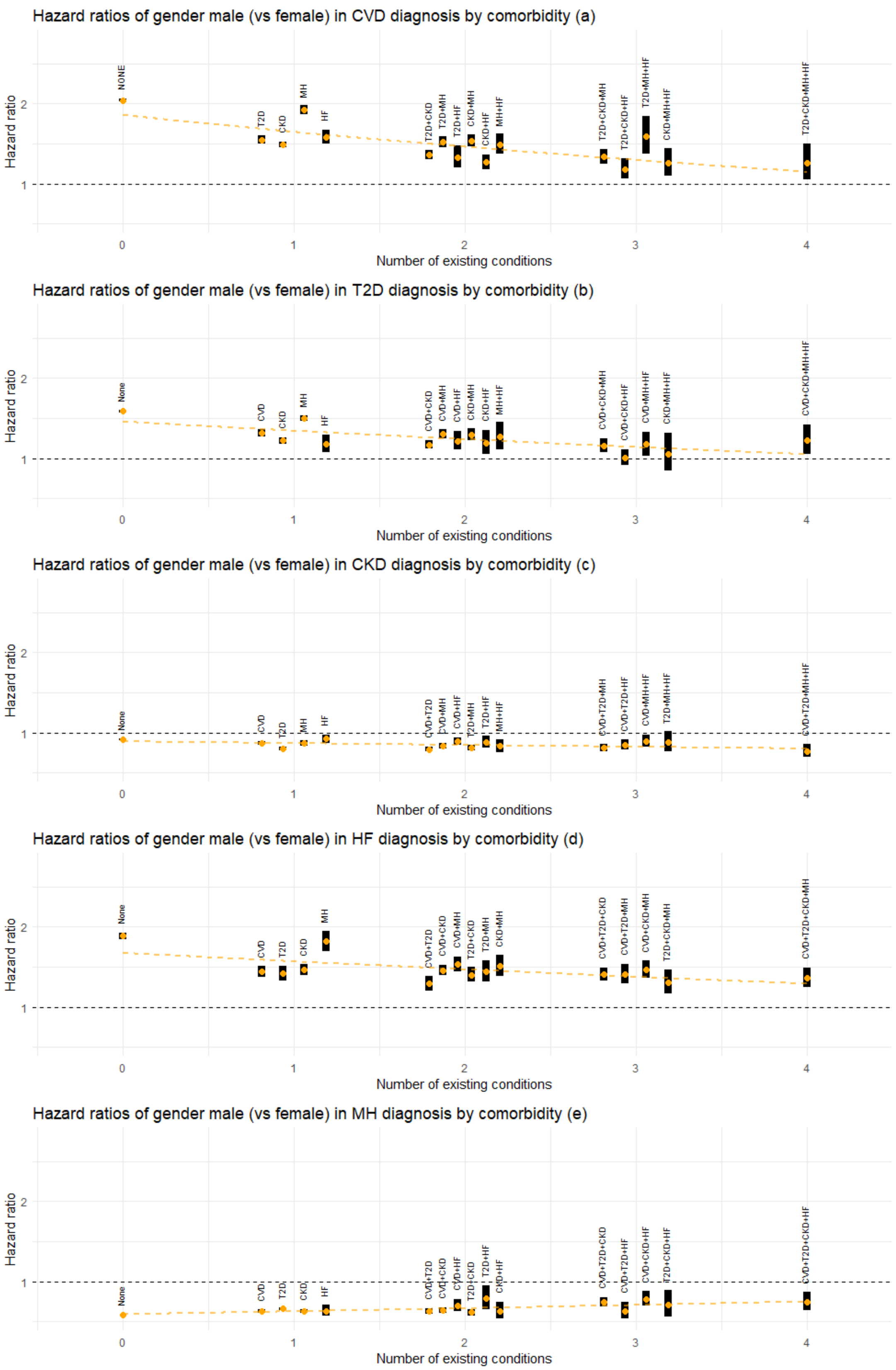


Figure H.3. Estimated association between gender (male) and the rate of disease transition by number of existing conditions and comorbidity status. Graphical settings are the same as in Figure 4 of the main paper. Abbreviations: CVD: cardiovascular disease; T2D: type-2 diabetes; CKD: chronic kidney disease; HF: heart failure; MH: mental health conditions.

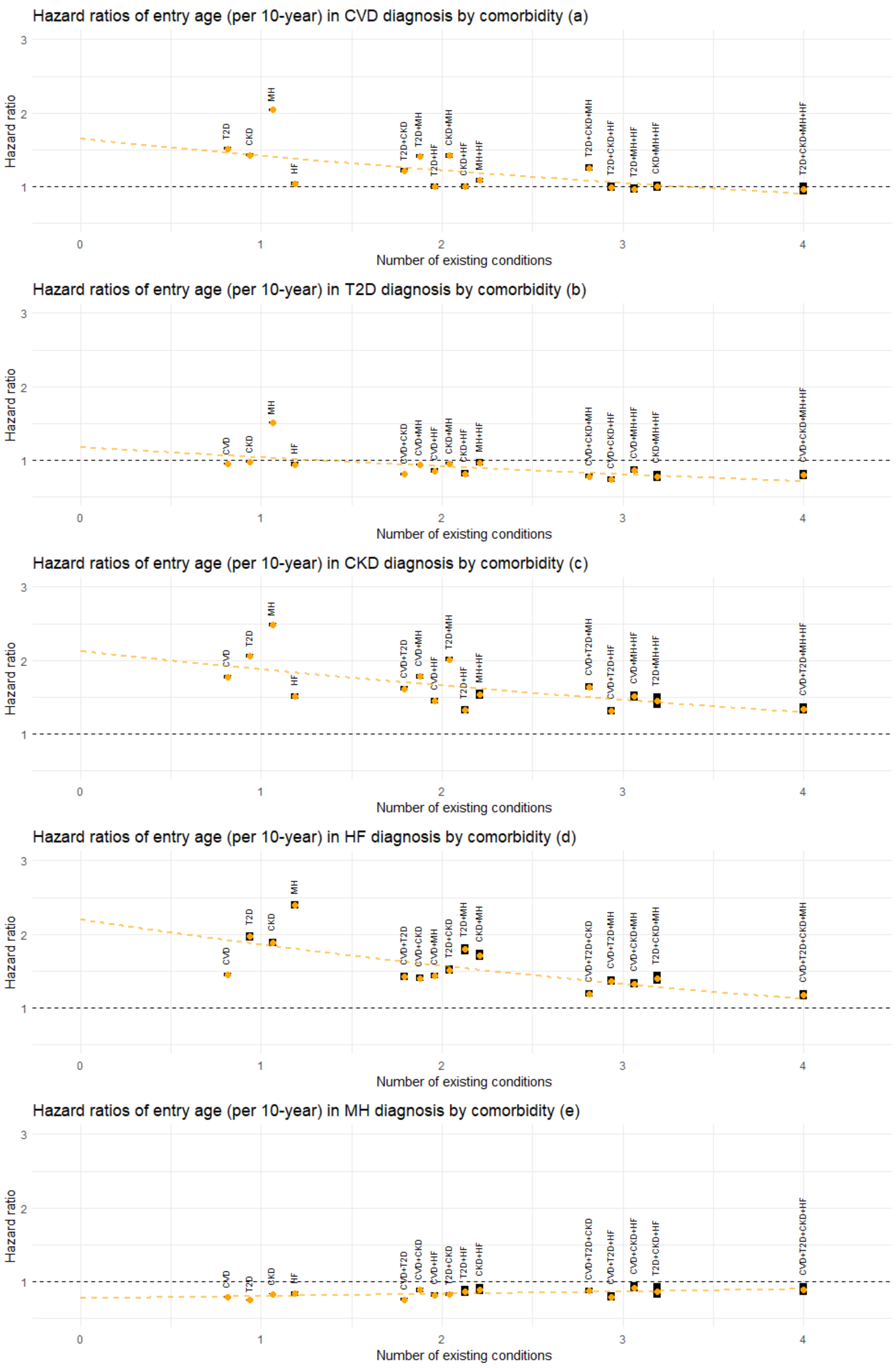


Figure H.4. Estimated association between age at entry to the current state (per 10 years) and the rate of next disease transition by number of existing conditions and comorbidity status. Graphical settings are the same as in Figure 5 of the main paper. Abbreviations: CVD: cardiovascular disease; T2D: type-2 diabetes; CKD: chronic kidney disease; HF: heart failure; MH: mental health conditions.
